# Whole-exome sequencing study identifies genes associated with Alzheimer’s disease and related dementias

**DOI:** 10.1101/2022.12.22.22283864

**Authors:** Xiaoyi Raymond Gao, Marion Chiariglione, Alexander J. Arch

**Affiliations:** Department of Ophthalmology and Visual Sciences, The Ohio State University, Columbus, OH 43210, USA; Department of Biomedical Informatics, The Ohio State University, Columbus, OH 43210, USA; Division of Human Genetics, The Ohio State University, Columbus, OH 43210, USA; Ohio State University Physicians Inc., Columbus, OH, USA

**Author notes:** Correspondence should be addressed to: Xiaoyi Raymond Gao, PhD, Department of Ophthalmology and Visual Sciences, Department of Biomedical Informatics, Division of Human Genetics, The Ohio State University, Columbus, OH 43212, USA.

## Abstract

Alzheimer’s disease (AD) is the most common form of late-onset neurodegenerative disease. Previous genome-wide association studies have identified numerous common genetic variants associated with AD. The contribution of rare variants to AD remains to be uncovered. AD-by-proxy, based on parental AD status, showed superior statistical power boost in recent AD studies. Using the UK Biobank (UKB) 368,865 whole-exome sequences of white British ancestry and AD-by-proxy (57,976 proxy AD cases and 310,889 non-AD proxy controls), we conducted the largest exome-wide association study of proxy AD to date. We identified 38 novel genes harboring rare variants for AD, such as *ANKRD36* (*P* = 6.02 × 10^−31^), *MMP13* (*P* = 1.08 × 10^−6^), *TUBA4A* (*P* = 7.15 × 10^−12^), and *ZNF296* (*P* = 4.19 × 10^−14^), demonstrating the power boost of aggregating rare variants and utilizing AD-by-proxy in gene discovery. We further replicated these genes in the FinnGen dataset. Notably, *MMP13* is a current drug target for AD and *TUBA4A* is a drug target for cognitive impairment in clinical trials. These results expand our knowledge of the genetic architecture of AD, especially the role of rare variants, and potential drug targets for AD.

## Introduction

Alzheimer’s disease (AD) is the most common form late-onset neurodegenerative disease. It affects millions of people worldwide^1^. The age for late-onset AD is typically 65 and older. The heritability of AD is about 60-80%^2^. Previous genome-wide association studies (GWAS) and meta-analyses have uncovered common variants for AD with 75 genetic loci identified to date^3^. However, the role of rare variants in AD remains to be discovered. Well established genes harboring rare variants with association to AD only include *TREM2, SORL1*, and *ABCA7*^4^. Studying rare variants that are associated with AD will help advance our understanding of the biological mechanisms and potential new therapeutic targets for this devastating disease.

Large-scale biobanks have propelled recent rare-variant genetic association studies with unprecedent novel discoveries^5-7^. For example, the UK Biobank (UKB), a study of half a million individuals living in the United Kingdom with deep genotyping and genomic data^8^, represents one of the most successful biobanks in biomedical research. It recently released whole-exome sequencies (WES) of 454,756 participants. A number of reports have used the UKB WES dataset for genetic association studies of quantitative traits and disease endpoints. However, for late-onset disease, e.g., AD, the number of cases can still be limited even in biobanks. To overcome this limitation, several recent studies used AD-by-proxy inferred from parental AD information and successfully made novel discoveries^3,9^.

The effectiveness of by-proxy phenotypes for GWAS was demonstrated by Liu and colleagues^10^. In a proxy case-control study design, proxy cases are the relatives of affected individuals^10^. Similarly, AD-by-proxy is to infer AD proxy case/control status based on parental AD history^9^. It is an approach that can significantly boost the effective sample size and statistical power of AD association tests^9,10^. The genetic correlation between AD-by-proxy and AD was found to be high (0.81)^9^. Hence, it is feasible to study AD through an AD-by-proxy approach. The robustness of this strategy in GWAS was also demonstrated recently^9^. AD-by-proxy provides an effective approach to conduct AD association studies utilizing large-scale biobank resources to obtain substantial gain in statistical power^9^.

In this study, we carried out an exome-wide association study (ExWAS) to identify genes associated with proxy AD using the UKB WES data. We further conducted replication of the identified genes in AD phenotypes in the FinnGen dataset, a biobank of over 200,000 Finnish people. To the best of our knowledge, this study represents the largest ExWAS of AD-by-proxy so far. Our results uncovered novel genes harboring rare variant with association to AD and advanced our understanding of the biological mechanisms of the disease. We further explored potential drug targets of the identified genes.

## Results

In this AD analysis using the AD-by-proxy phenotype, a total of 368,865 participants of white British ancestry (57,976 cases and 310,889 controls) from UKB were included following Jansen et al.’s proxy AD definition^9^. The mean (standard deviation) age was 59.2 (6.9) years for the cases and 56.7 (8.1) years for the controls. Among the AD cases, 33,128 (57.1%) were female, and among the non-AD controls, 172,152 (55.4%) were female.

In this ExWAS analysis, we replicated 26 previously known AD genes, including all established genes harboring rare variants (*TREM2, SORL1*, and *ABCA7*) and identified 38 novel potential AD-by-proxy genes (*P* < 2.5 × 10^−6^) from our gene-based analysis using SAIGE^11^, a state-of-the-art tool for gene-based association testing. The genomic control lambda was 1.0, indicating our analysis is properly controlled for population structure and cryptic relatedness. The corresponding quantile-quantile plot is shown in Supplementary Figure 1. Table 1 shows the list of gene-based associations with AD-by-proxy. Genes were considered novel if they were not found associated with AD within the GWAS catalog or the Alzheimer’s Disease Variant Portal (ADVP) databases (see web resources). *ANKRD36* shows a highly significant association with proxy AD (*P* = 6.02 × 10^−31^), followed by other hits, such as *PRB4* (*P* = 7.78 × 10^−15^), *ZNF296* (*P* = 4.19 × 10^−14^), *TTK* (2.10 × 10^−12^), *TUBA4A* (*P* = 7.15 × 10^−12^), *TUBA4B* (*P* = 7.98 × 10^−12^), and *MMP13* (*P* = 1.08 × 10^−6^). We further queried the FinnGen website for the association of the identified genes with AD. Many of these genes were identified to have strong associations with AD in FinnGen. For example, there is a highly significant association with AD for *ZNF296* (*P* = 8.9 × 10^−100^), *ADAM8* (*P* = 2.3 × 10^−6^), *HIGD1B* (*P* = 6.6 × 10^−6^), *EFTUD2* (*P* = 6.6 × 10^−6^) and *TTK* (*P* = 1.4 × 10^−5^). A Manhattan plot of the gene-based p-values is shown in Figure 1.

**Table 1.**
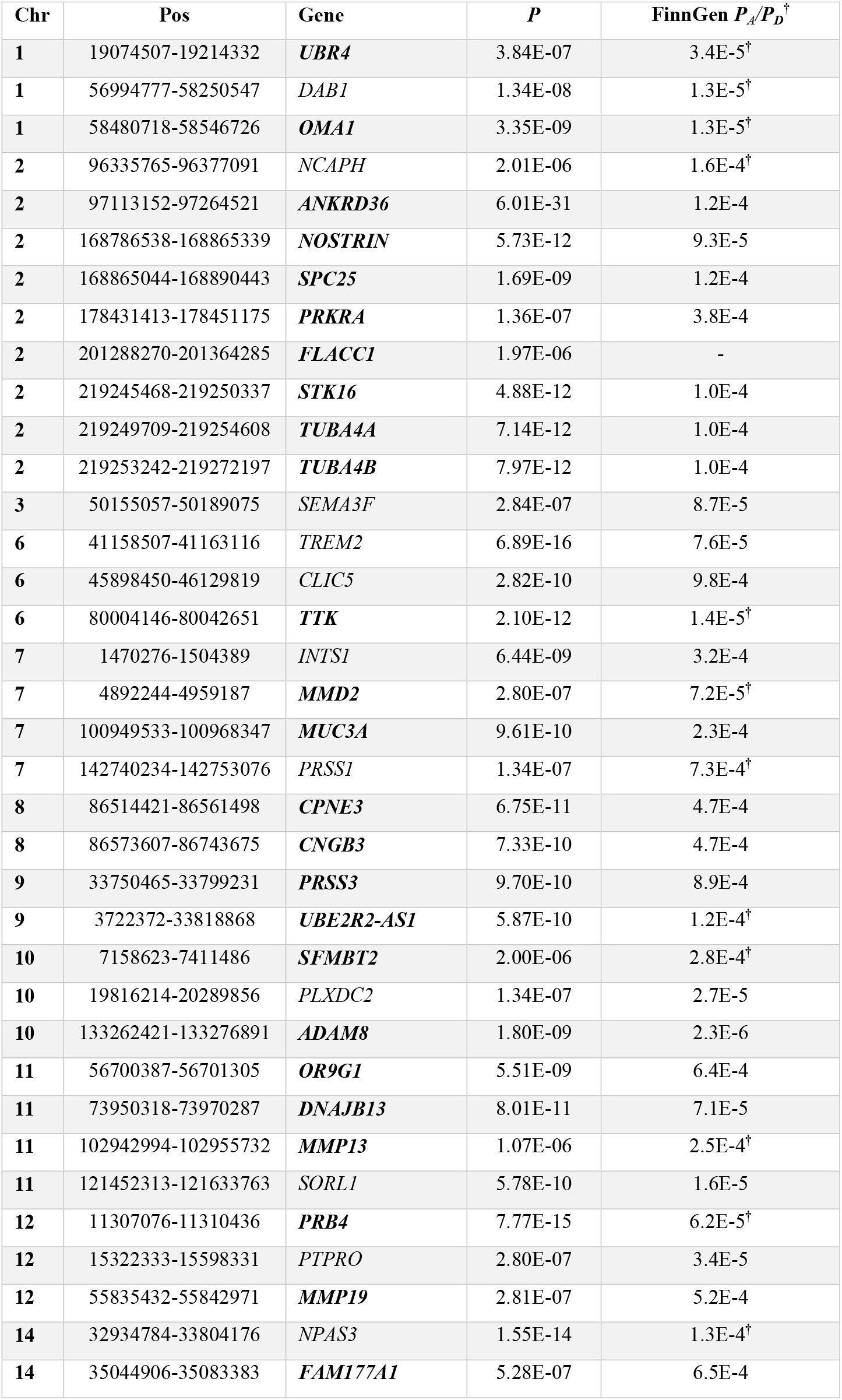
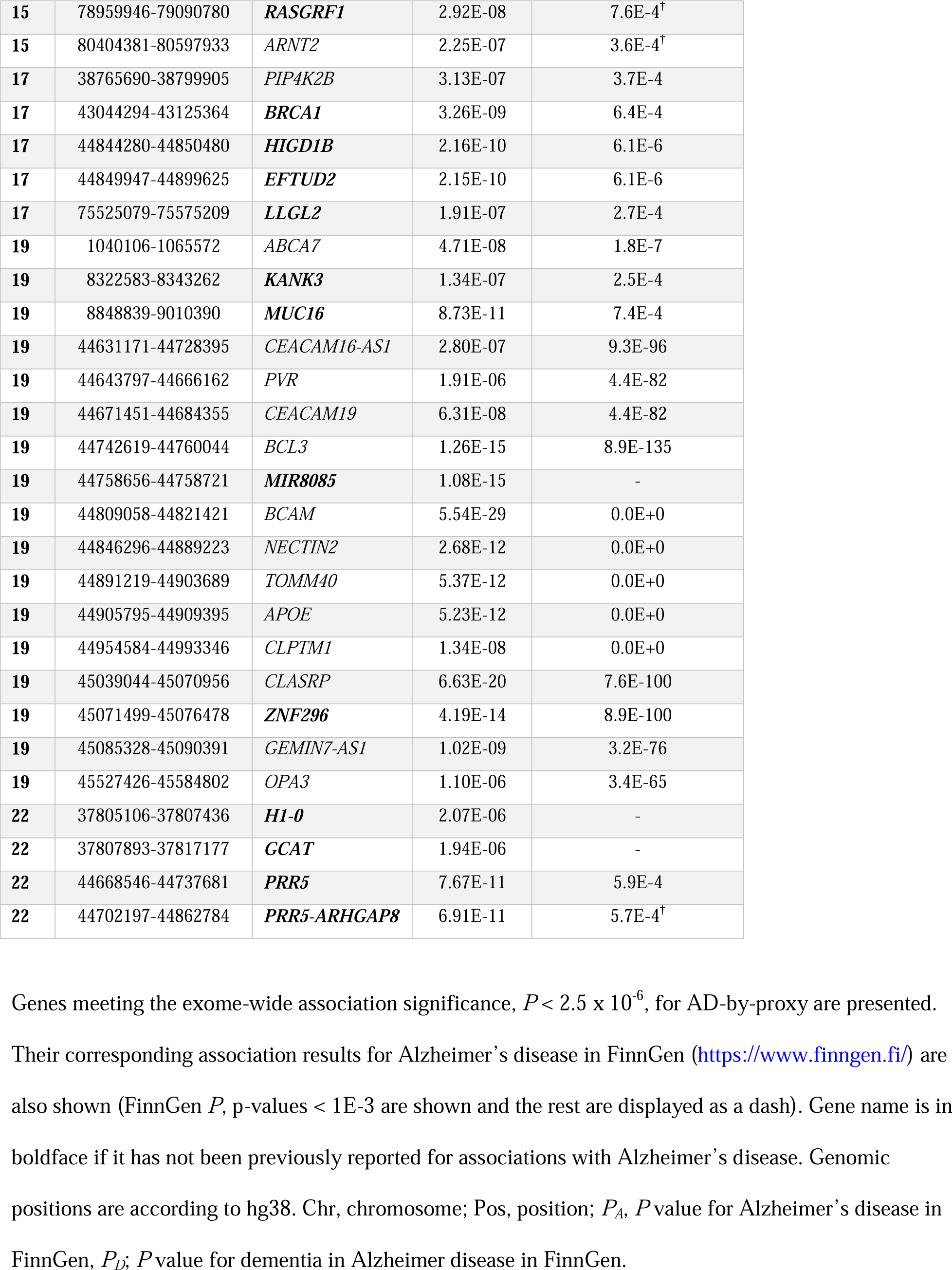
Exome-wide significant gene-based results for proxy AD.

**Figure 1.**
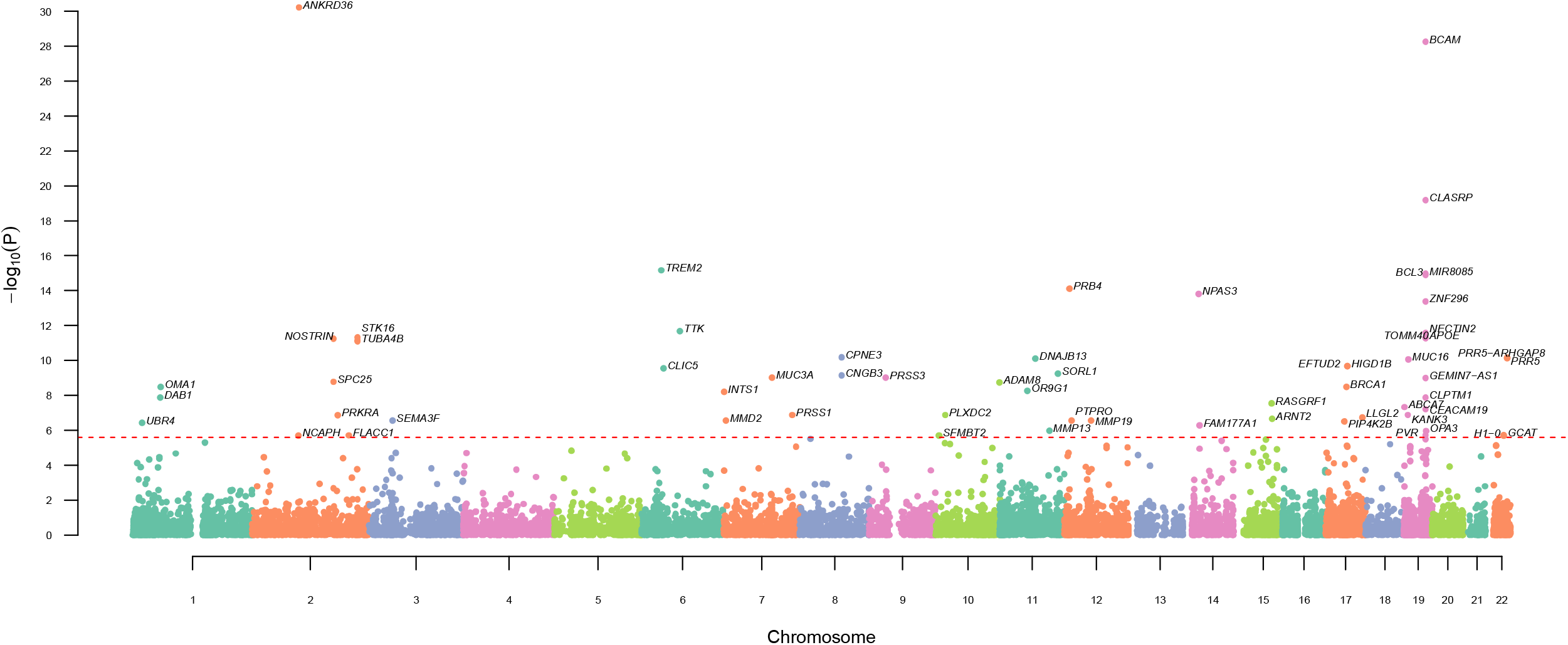
Manhattan plots displaying the -log_10_(*P*) for the association of gene-based test results. Gene-based results showing the association strength between proxy AD and genes across the genome. The dotted horizontal line represents the exome-wide association significance level, *P* < 2.5 × 10^−6^. To aid visualization, chromosomes are delimitated by colors. Genomic positions are according to hg38.

To seek biological support for the identified genes, we used the gene expression and differential expression information in the Genevestigator^12^ and the Agora databases, respectively. Supplementary Figure 2 displays the bulk tissue gene expression information from Genevestigator. Most of the identified novel genes, such as *ANKRD36, TUBA4A, TUBA4B, MMP13, MUC16, TTK*, and *ZNF296*, are expressed in the brain. Figure 2 shows the differential expression (DE) evidence for these identified genes from the Agora knowledge portal, a platform for exploring AD genes, including DE, initially developed by the National Institute on Aging’s Accelerating Medicines Partnership in Alzheimer’s Disease (AMP-AD) consortium. In the figure, the diameter and the color of the circles indicate p-value and log2 fold change, respectively. Many of the significant DE genes are perturbed in parahippocampal gyrus and temporal cortex. Several of identified genes, such as *RASGRF1, HIGD1B*, and *LLGL2*, show significant DE across most of the AD-related brain regions.

**Figure 2.**
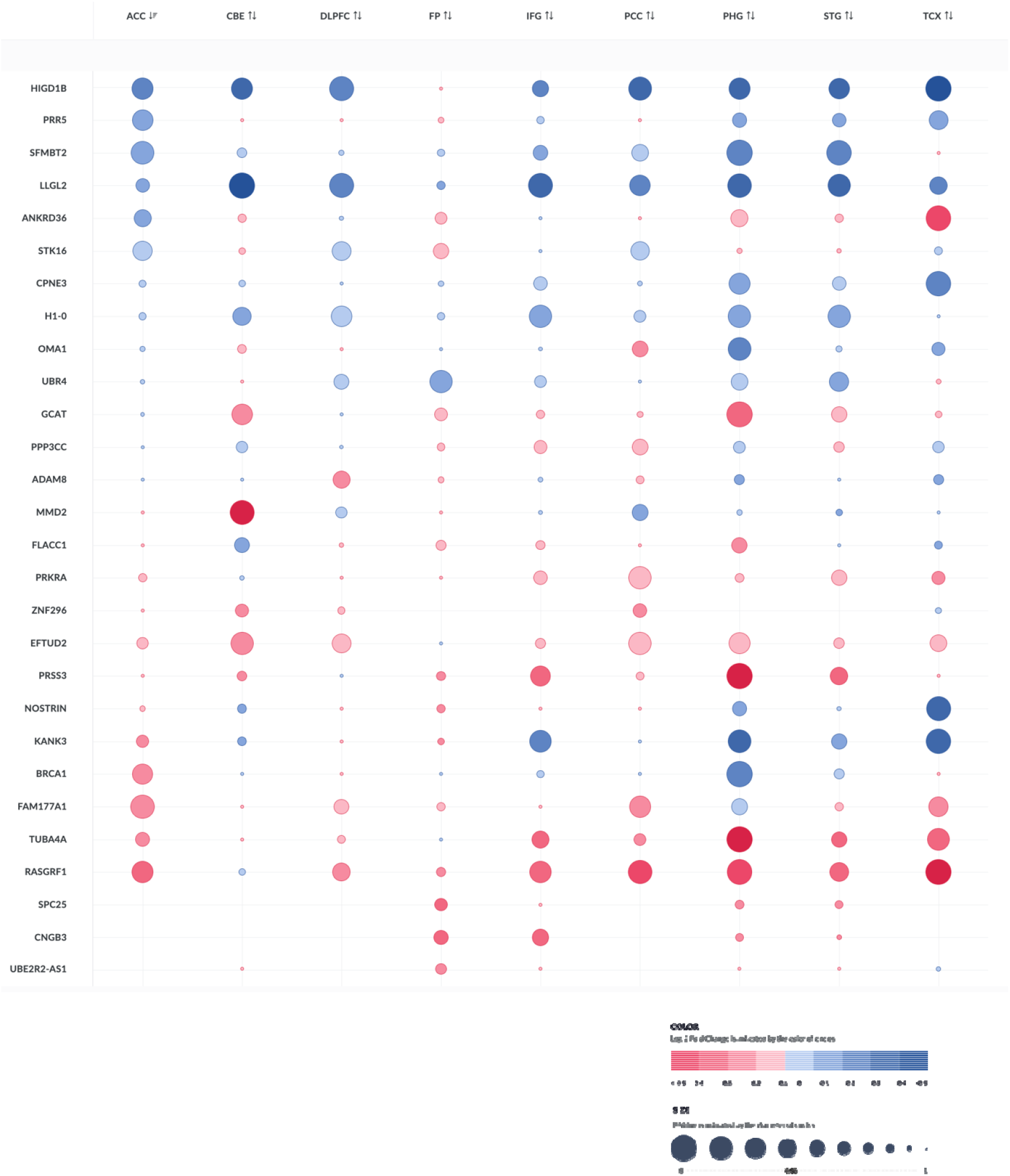
Differential expression evidence from Agora. Differential expression results were queried from the Agora website. In the figure, the diameter and the color of the circles indicate p-value and log2 fold change, respectively. Abbreviations: ACC, anterior cingulate cortex; CBE, cerebellum; DLPFL, dorsolateral prefrontal cortex; FP, frontal pole; IFG, inferior frontal gyrus; PCC, posterior cingulate cortex; PHG, parahippocampal gyrus; STG, superior temporal gyrus; TCX, temporal cortex.

The novel genes also included some promising drug targets for AD. To query potential drug targets, we used the online Open Targets tool. Several of the identified genes associated with proxy AD have known drug targets in clinical trials, as shown in Table 2. The genes *TUBA4A* is targeted by Davunetide, a tubulin stabilizer, which may serve as a possible treatment for cognitive impairment. The drug Doxycycline, a matrix metalloproteinase inhibitor, targets the gene *MMP13* and is being researched for use against AD. *MUC16* and *TTK* are targeted by Oregovomab and BAY-1217389 for treating ovarian cancer and cancer, respectively.

**Table 2.** Known drug targets for the identified genes associated with Alzheimer’s.

| Gene | Known Drugs | Mechanism of Action | Disease Information |
| --- | --- | --- | --- |
| ***TUBA4A*\*** | Davunetide | Tubulin stabiliser | Cognitive impairment<br>Progressive supranuclear palsy<br>Schizophrenia |
|  | Sagopilone | Tubulin stabiliser | Central Nervous System Neoplasm<br>Glioblastoma multiforme |
|  | Lorvotuzumab mertansine | Tubulin inhibitor | Malignant peripheral nerve sheath tumor<br>Glioblastoma multiforme |
|  | ANG1005 | Tubulin inhibitor | Malignant glioma<br>Brain neoplasm<br>Neoplasm |
|  | Verubulin | Tubulin disrupting agent | Glioblastoma multiforme<br>Brain neoplasm<br>Neoplasm |
|  | Lexibulin | Tubulin inhibitor | Glioblastoma multiforme<br>Multiple myeloma |
| ***MMP13*\*** | Doxycycline | Matrix metalloproteinase 13 inhibitor | Alzheimer disease<br>Brain injury |
|  | Doxycycline Anhydrous | Matrix metalloproteinase 13 inhibitor | Alzheimer disease |
| ***MUC16*** | Oregovomab | Mucin-16 other | Ovarian cancer<br>Pancreatic adenocarcinoma<br>Peritoneum cancer<br>Ovarian neoplasm<br>Fallopian tube cancer<br>Ovarian carcinoma |
|  | Abagovomab | Mucin-16 other | Ovarian cancer<br>Peritoneum cancer<br>Fallopian tube cancer |
|  | Sofituzumab Vedotin | Mucin-16 binding agent | Pancreatic carcinoma<br>Ovarian cancer |
| ***TTK*** | BAY-1217389 | Dual specificity protein kinase TTK inhibitor | Cancer |
|  | BAY-1217389 | Dual specificity protein kinase TTK inhibitor | Cancer |

The table shows existing drugs that target the rare-variant genes identified.
\*These lists are not all inclusive. *TUBA4A* has a total of 19 drugs with 69 indications and *MMP13* has a total of 4 drugs with 103 indications.

## Discussion

In this study, we conducted the largest ExWAS of proxy AD to date using the WES data from UKB. By utilizing an AD-by-proxy approach, we have expanded our knowledge of the role of rare variants for AD and related dementias. We identified 38 novel genes for proxy AD, demonstrating the power of AD-by-proxy and rare-variant aggregation in gene discovery. We also replicated 26 previously reported AD genes, including all the well-established genes harboring rare variants, i.e., *TREM2, SORL1*, and *ABCA7*, further demonstrating the robustness of our approach. Of note, among the novel genes, *MMP13* and *TUBA4A*, have already been targeted in clinical trials for AD and cognitive impairment.

AD gene mapping can be hindered by the limited number of cases available, especially for rare-variant investigation. Previous approaches have used meta-analysis, which has been successful in identifying numerous common variants associated with AD. Another approach is to use AD-by-proxy^9^ taking advantage of the enormous resources available in large-scale biobanks, e.g., UKB. This disease-by-proxy approach has been used in several recent studies on GWAS datasets^3,9,13^. Here, we demonstrated its effectiveness using the UKB WES data and identified novel genes for AD.

The identified ExWAS significant genes show many interesting patterns. The most significant one, *ANKRD36* (*P* = 6.01 × 10^−31^), is highly significantly associated with autoimmune disease in FinnGen (Beta 8, *P* = 9.6 × 10^−10^). A recent report showed that AD may be rooted in an autoimmune response^14^. *ANKRD36* has also been reported for an association with schizophrenia^15^, a severe psychiatric illness that may have a white matter deficit connection with AD^16^. *ANKRD36* is also involved in hypertension^17^, a risk factor for AD. Many genes, such as *CEACAM19, BCL3, BCAM, NECTIN2, TOMM40, APOE, CLASRP*, and *OPA3*, are in the about 1M region around *APOE* on chromosome 19 and show both significant common- and rare-variant associations with AD. We further identified two other genes, i.e., *MIR8085* (*P* = 1.09 × 10^−15^) and *ZNF296* (*P* = 4.19 × 10^−14^), that showed significant rare-variant associations with proxy AD in the 1M region around *APOE*. Many of the other significant genes, such as *DAB1, OMA1, PVR, CEACAM19, BCL3, CLASRP*, and *ZNF296*, are highly significantly associated with statin medication in FinnGen (online Beta 8 results), which may indicate the interrelationship between AD and cardiovascular health.

There is a dearth of drugs available for AD. With the goal of conquering AD by 2025^18^, we clearly need more candidate genes for AD drug targets. Genes in our list has already given promising results. For example, *MMP13* (*P* = 1.07 × 10^−6^) in our novel gene list is an AD drug target for Doxycycline and has a total of 4 drugs with 103 indications. *TUBA4A* is in clinical trials for cognitive impairment and has a total of 19 drugs with 69 indications. *MUC16* and *TTK* are known targets for some cancer drugs, which may possibly be repurposed for AD. The other genes in our list may serve as novel drug targets as well.

Our study is not without limitations. AD-by-proxy is a proxy for AD based on parental AD history and is less defined than clinically diagnosed AD. However, due the high heritability of AD, the case status for offspring of AD parents is expected to be high. Coupled with large sample size of biobanks, like UKB, proxy AD can substantially increase the statistical power of AD discoveries. Moreover, AD-by-proxy has been proven successful in AD discoveries in previous AD GWAS studies^3,9^. Using this approach, we not only identified novel AD genes but also confirmed 26 known ones. Most of the novel ones showed DE for AD in the Agora resources. Several of these novel genes identified are also drug targets for AD in clinical trials, which further demonstrated the effectiveness of our AD-by-proxy approach in AD genetic discoveries. Nevertheless, replication of our results in clinical diagnosed AD in other datasets and diverse populations is needed. There are many different ways to analyze rare variants, e.g,, including synonymous variants^19^, sliding window^20^, and all inclusive^21^. To our knowledge, there is no single rare-variant analysis method that performs the best in all situations. Using other approaches may identify more novel AD genes.

In conclusion, we carried out the largest ExWAS of AD-by-proxy to date. In addition to improving our understanding of the genetic architecture of AD and related dementias, especially the role of rare variants, the results of our study may serve as a valuable resource for exploring potential drug targets for AD given the relevance of *MMP13* and *TUBA4A*, all of which are in our novel gene list, as drug targets for AD and cognitive impairment.

## Materials and Methods

### UKB Resource

UKB is a large prospective cohort study comprising half a million participants living in the United Kingdom. The details of the UKB cohort have been previously reported. Briefly, participants aged 40 to 70 years and registered with the National Health Services were enrolled during the baseline recruitment from 2006 to 2010. Family history, lifestyle information, self-reported health information, electronic medical records, and DNA samples were collected. Approximately 94% of the participants reported their ethnic background as white, and we used individuals of white British ancestry for this study. Our access to the UKB data was obtained under application number 23424, and we obtained fully de-identified data.

### FinnGen Resource

FinnGen is a large Finnish biobank that focuses on the Finnish population. It has enrolled over 200,000 participants, who are aged 18 or older and have a median age of 63^22^. By the end of 2023, it is projected to have more than half a million participants^22^. Comprehensive genotype and phenotype data have been generated and collected since 2017. Genotyping was done using the custom Axiom FinnGen1 array as well as legacy arrays. The genotypes were further imputed to 17 million markers based on whole-genome sequences of Finns. Phenotypes were primarily constructed using International Classification of Disease Ninth and Tenth Revision (ICD-9 and ICD-10) codes. FinnGen GWAS summary statistics for various phenotypes are publicly available (see web resources).

### UKB WES and Quality Control

The WES data for UKB participants were generated at the Regeneron Genetic Center^5,23^. Previously, details on the sequencing, variant calling, and quality control processes have been reported^6,23^. In brief, the Illumina NovaSeq 6000 platform was used for sequencing with 75 base pair paired-end reads, and the SPB protocol was utilized for variant calling and quality control^24^. The WES data was of high quality, with over 20x coverage of 95.8% of targeted bases. We overlaid the data with our derived proxy AD phenotype and retained samples with missing rates less than 2.5% and autosomal variants with minor allele frequencies (MAFs) less than 0.01, call rates greater than 95%, and minor allele counts greater than or equal to 1 (19.6 million rare variants). We used VEP^25^ and annovar^26^ for annotating the variants.

### AD-by-proxy phenotype in UKB

We derived the AD-by-proxy phenotype following Jansen *et al*.’s previous report on proxy AD^9^ based on the AD information obtained through the intake UKB questionnaire and electronic health records. “By proxy” indicates that the phenotype built includes information about the status of the immediate biological family of the individual regarding the illness studied. The phenotype also includes the status of the individual itself regarding whether they have AD or not. During the intake process with UKB, each individual was asked about their biological mother’s and father’s illnesses and whether they suffer from AD (data fields 20107 and 20110 for the illnesses of father and mother, respectively) as well as the current parent’s age (data fields 2946 and 1845) or age when they passed away if applicable (data fields 1807 and 3526). Based on these fields, if an individual’s parent was found to have AD he/she was given a “risk” score of 1, on the other hand, if they were found to be unaffected, their score was computed using their age or age at death to consider the fact that they might not have been old enough for a diagnosis of this late-onset disease. The formula used for this is the proportion of the parent’s age to age 100: *parental_risk_score* (100 - *age*) / 100. This parental risk score for the unaffected parent, was also capped at 0.32, corresponding to a risk similar to that of the general occurrence of AD in the population^27^. We used the International Classification of Disease Tenth Revision (ICD-10) codes (data fields 41202 and 41204 for diagnoses - main and secondary, respectively) to characterize a participant’s own illnesses information on AD. Participants with the codes G30 (AD) or F00 (Dementia in AD) were considered cases and were given the maximum “risk” score possible of 2 regardless of the AD status of their parents. The resulting AD-by-proxy score (including biological parents’ information) is between 0 and 2 with 0 representing both parents being unaffected and 2 when both parents or the individual were affected. Anyone with a score over 1 was considered a case. We also performed quality control on this phenotype and removed individuals who answered “do not know” or “prefer not to answer” on the questionnaires about their parents’ illnesses, individuals without any parents’ age, outliers for heterozygosity, individuals with ten or more third-degree relatives, and individuals with sex chromosome aneuploidy. For this study, we restricted our analysis to self-identified white British (UKB data field 21000) participants and removed outliers with genetic ancestry at least six standard deviations from the means of the first two principal components. This resulted in 57,976 AD cases and 310,889 non-AD controls (white participants) after overlapping with the WES data.

### Gene-based ExWAS analysis

We used SAIGE^11^, a generalized mixed model approach that can adjust for both genetic relationship and population stratification, to carry out our gene-based association tests. SAIGE includes several rare variant collapsing and aggregation tests, such as SKAT-O^28^, burden^29^, and SKAT^30,31^. We used the SKAT-O approach, which adaptively combines the strengths of both the burden and SKAT test statistics and demonstrates advantages in statistical power and robustness^32^. We included predicted loss of function (pLOF) variants, such as stop gained, stop lost, start lost, splice donor, splice acceptor, and frameshift, based on the VEP^25^ annotation and gnomAD pLOF variants^33^, as well as missense variants (MAF < 0.005). We adjusted for age, sex, and the first 10 principal components of genetic ancestry. Genes with P < 2.5 × 10^−6^ were declared ExWAS significant.

### AD lookup in FinnGen

To replicate our significant genes, we queried the FinnGen^22^ online GWAS summary statistics (Beta 8, https://www.finngen.fi/) and checked their associations with AD phenotypes in the FinnGen database. There are 14 endpoint AD phenotypes in FinnGen. We queried the Alzheimer’s disease (wide definition), which is defined as F00*, G30 (ICD10 hospital discharge or cause of death), 3310 (ICD9 hospital discharge or cause of death), 29010 (ICD8 hospital discharge or cause of death), 307 (KELA codes), and N06D (ATC codes), and dementia in Alzheimer disease, which is defined as F00* (ICD10 hospital discharge or cause of death) and 29010 (ICD8 hospital discharge or cause of death) based on the FinnGen health registries (https://risteys.finngen.fi/).

### Gene Expression Evidences

To query bulk RNA gene expression information in brain tissues, we used Genevestigator^12^, an online gene expression database. Expression profiles were obtained from 2,041 human nervous system tissues and displayed as box plots indicating low, medium, or high expression for the corresponding specific tissue. To query differentially expressed genes, we used Agora (see web resources), a web-based application that hosts evidence for whether or not genes are associated with AD.

### Drug Targets Prioritization

To identify potential drug targets for the genes identified in our analyses, we used the Open Targets^34^ online resource. We queried the website for known drugs, mechanisms of action (source ChEMBL), and disease information related to AD. This drug information provides valuable insights into the relevance of the identified genes to AD and potential drugs for repurposing.

## Data Availability

All data produced in the present work are contained in the manuscript.

## Acknowledgements

This work was supported in part by grants from the National Institute on Aging, National Institutes of Health (NIH; Bethesda, MD, USA): RF1AG060472. The content is solely the responsibility of the authors and does not necessarily represent the official views of the NIH. We would like to thank the study participants and investigators from the UK Biobank as well as the staff who aided in data collection and processing. We also want to acknowledge the participants and investigators of the FinnGen study.

## Conflict of Interest Statement

None declared.

### Web Resources

The URLs for downloaded data and programs:

Agora, https://agora.adknowledgeportal.org

Alzheimer’s Disease Variant Portal (ADVP), https://advp.niagads.org

ANNOVAR, http://annovar.openbioinformatics.org/

ChEMBL, https://www.ebi.ac.uk/chembl/

GWAS Catalog, https://www.ebi.ac.uk/gwas/

FinnGen, https://www.finngen.fi/

SAIGE, https://github.com/weizhouUMICH/SAIGE

UK Biobank, https://www.ukbiobank.ac.uk

VEP, https://useast.ensembl.org/info/docs/tools/vep/index.html

R, https://www.r-project.org

