## Supplementary Information for "Whole-exome sequencing study identifies genes associated with Alzheimer’s disease and related dementias"

**Supplementary Figure 1. Quantile-quantile plots for gene-based results.**

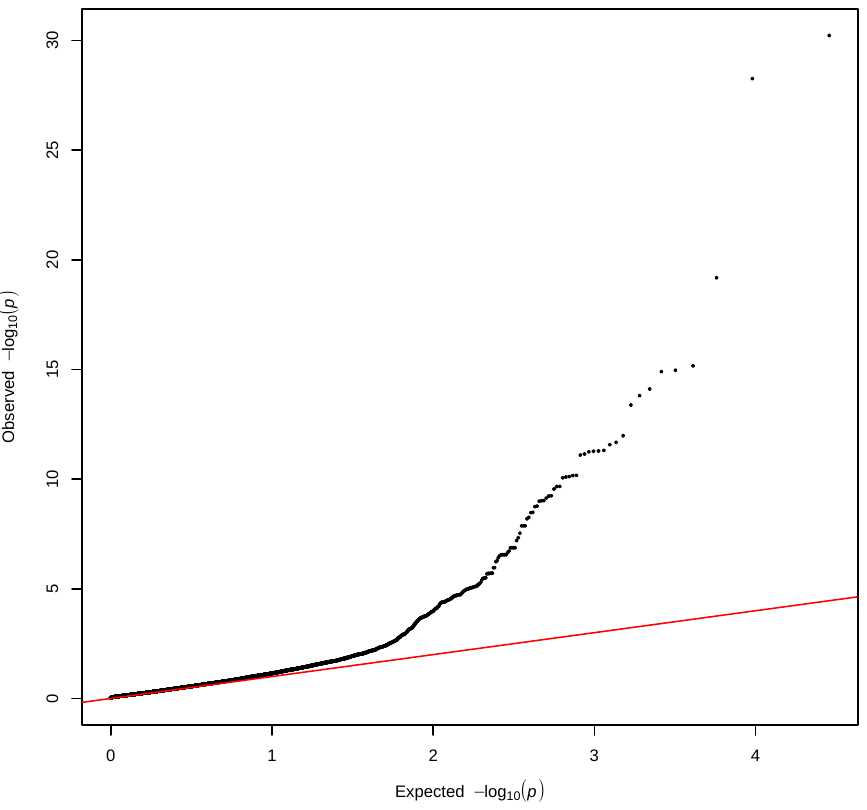

**Supplementary Figure 2. Gene expression profiles from Genevestigator.**

Gene expression profiles were generated with GENEVESTIGATOR^®^ (source: Hruz T*, et al.* Genevestigator v3: a reference expression database for the meta-analysis of transcriptomes. *Adv Bioinformatics* **2008**, 420747 (2008). https://doi.org/10.1155/2008/420747). Genes are listed alphabetically according to gene name.

***ADAM8***
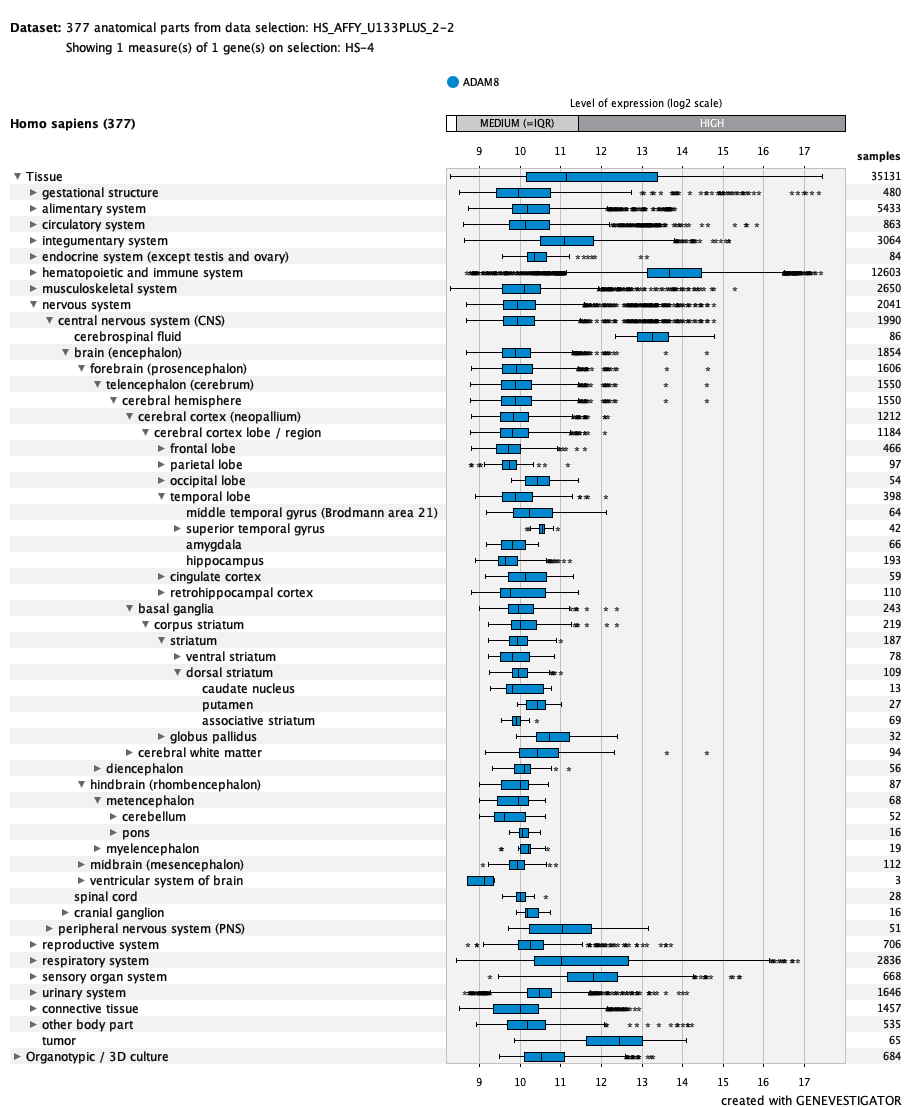

***ANKNRD36***
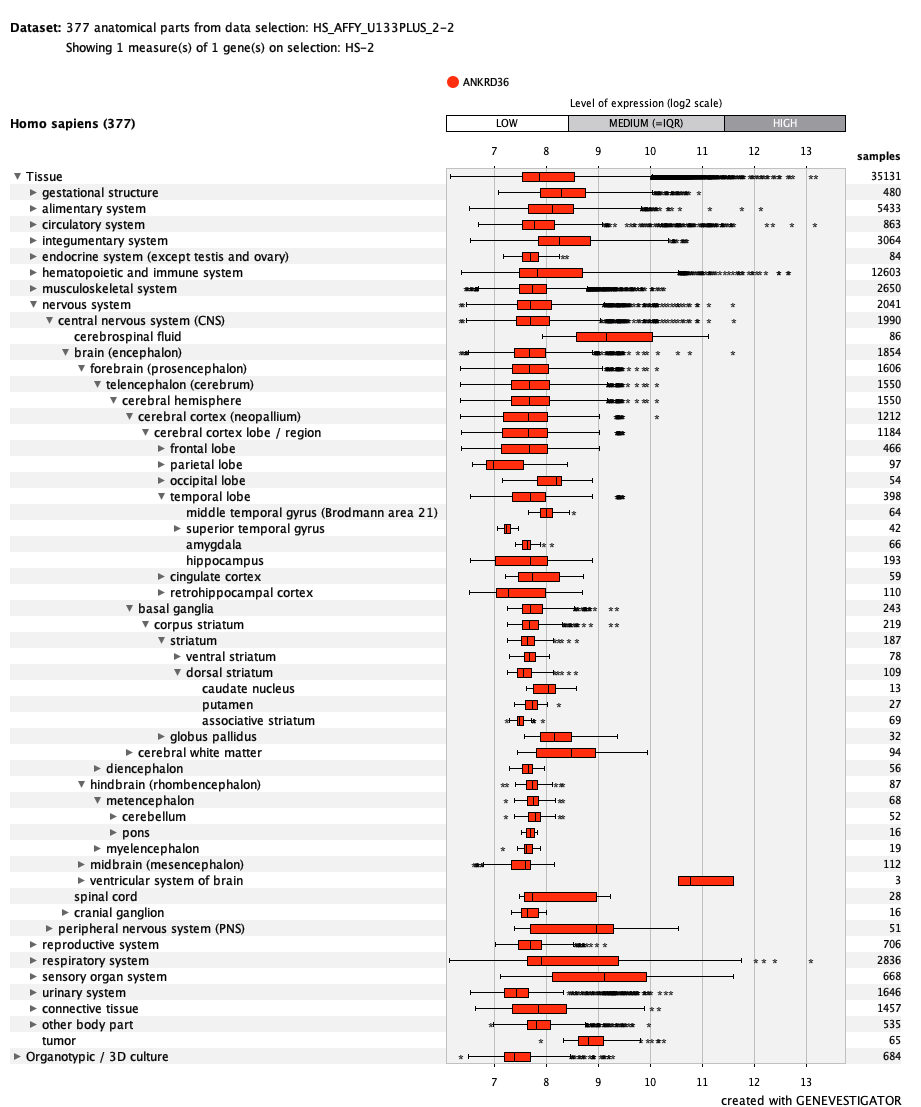

***BRCA1***
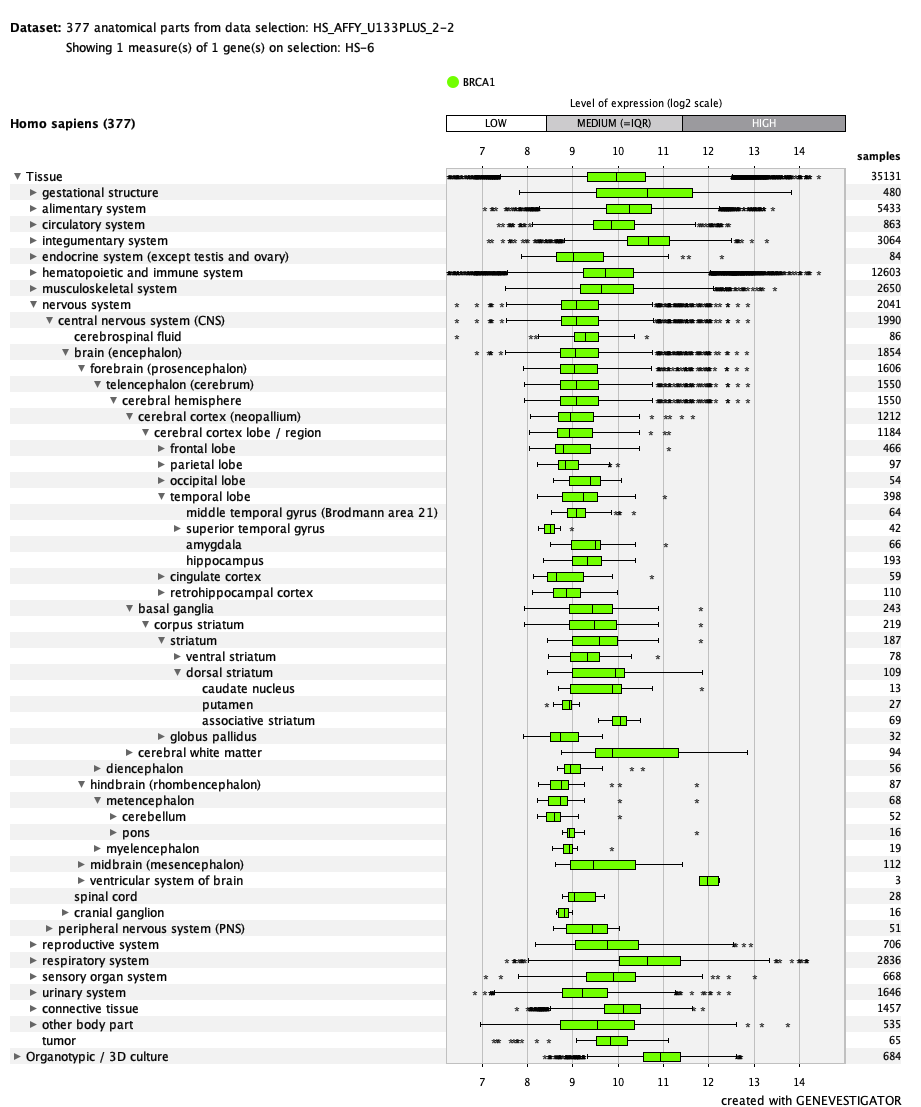

***CNGB3***
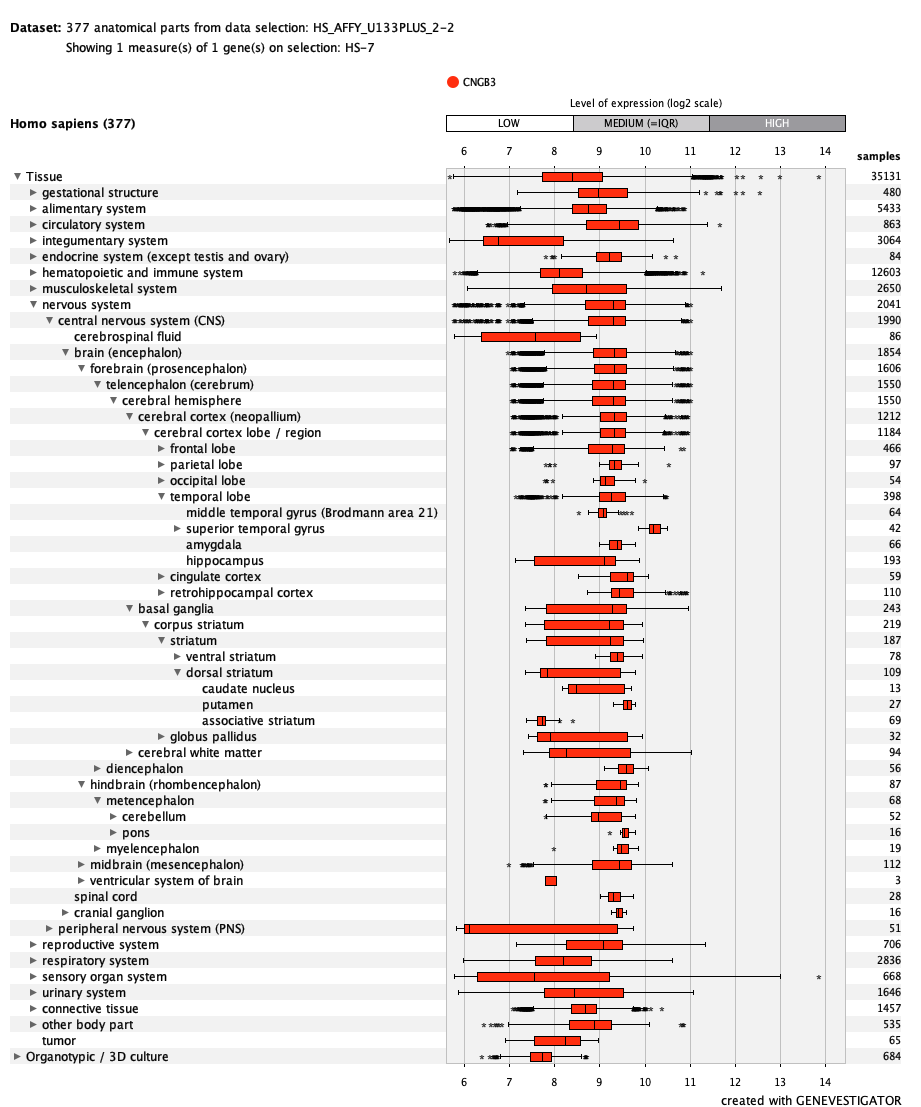

***CPNE3***
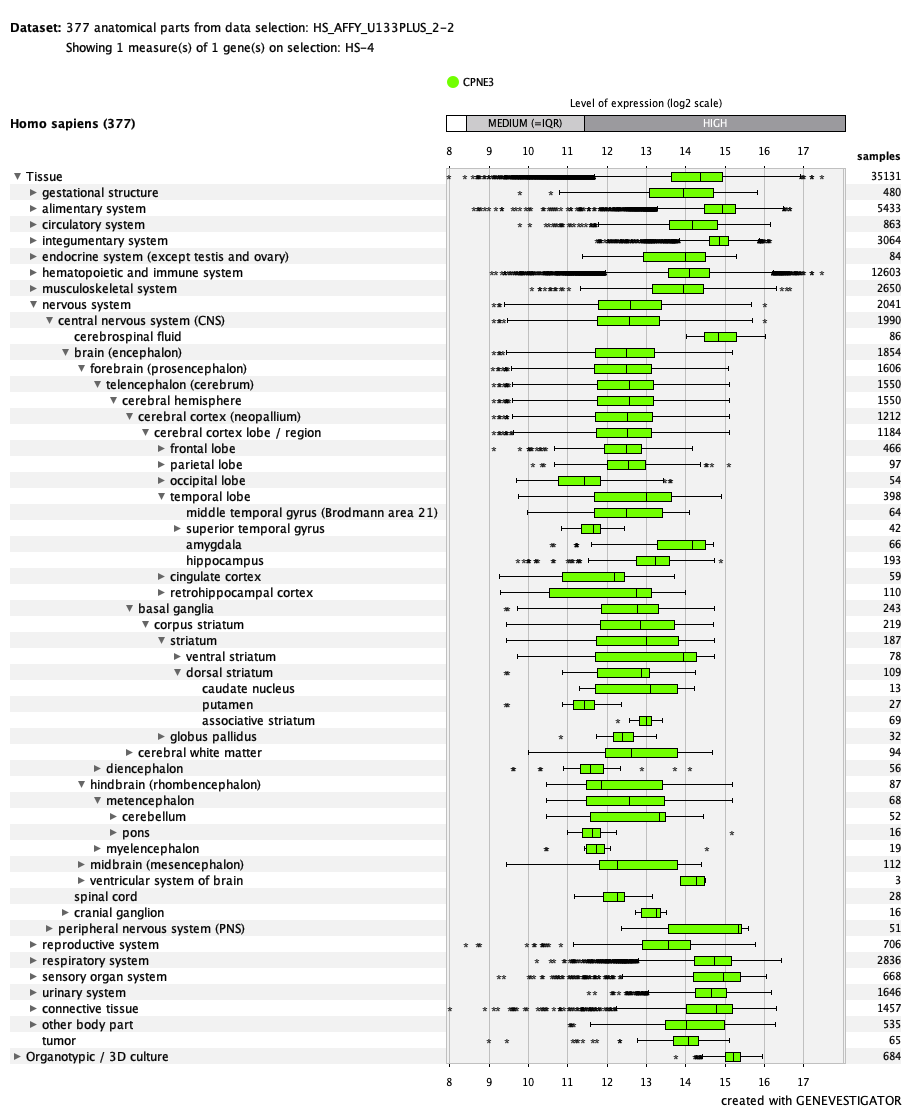

***DNAJB13***
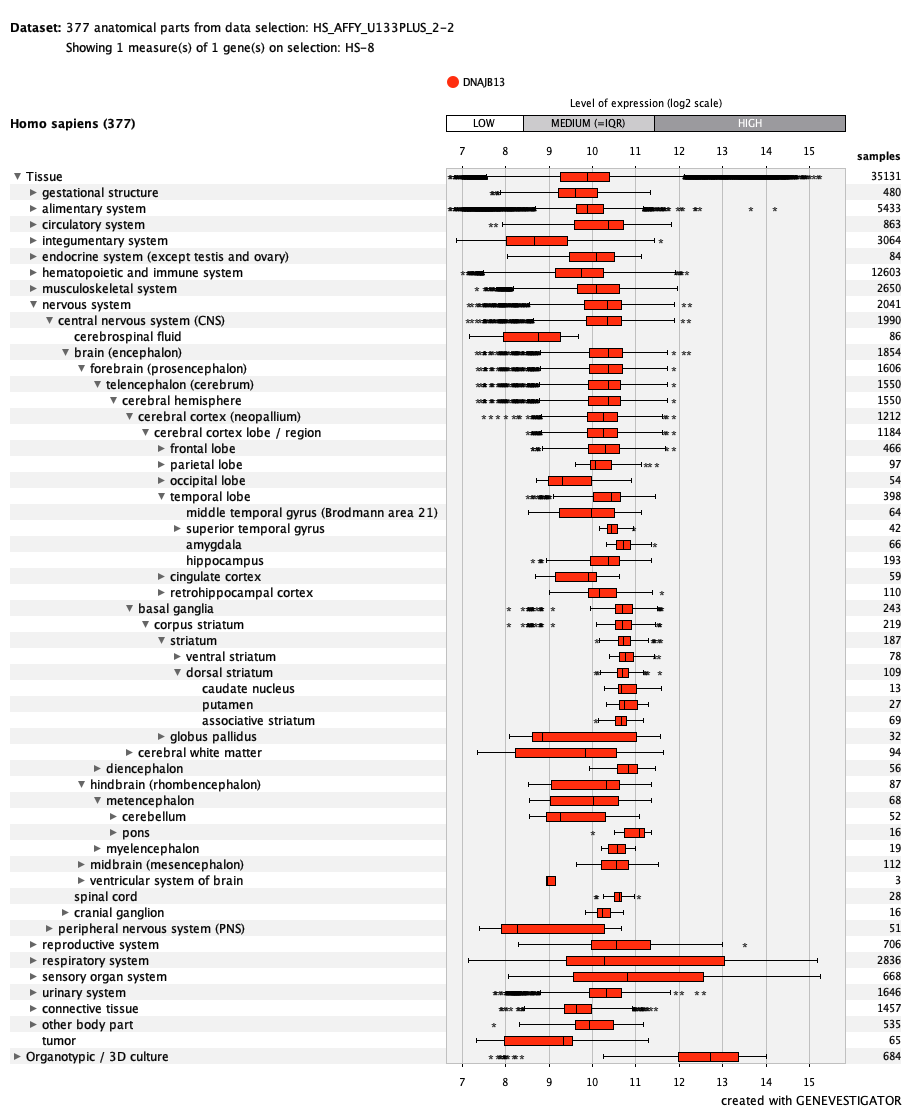

***EFTUD2\***
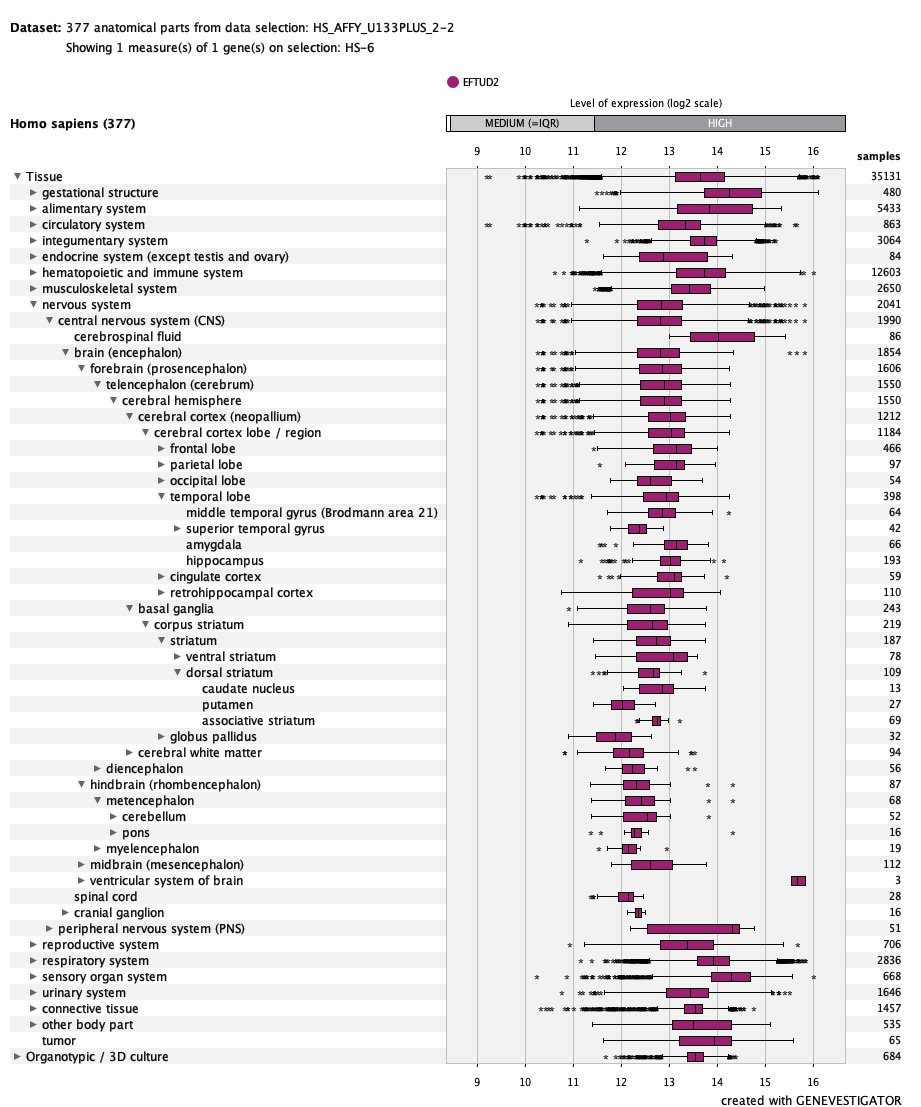

***FAM177A1***
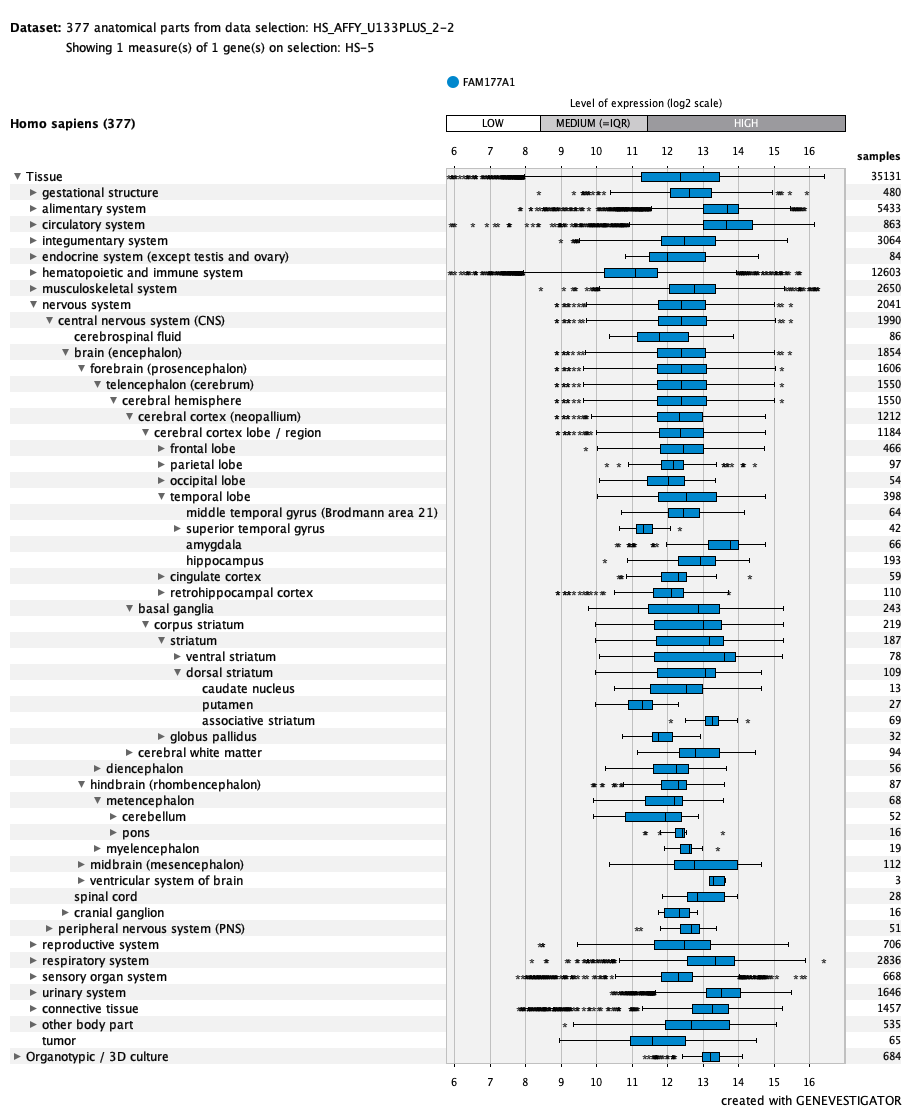

***FLACC1***
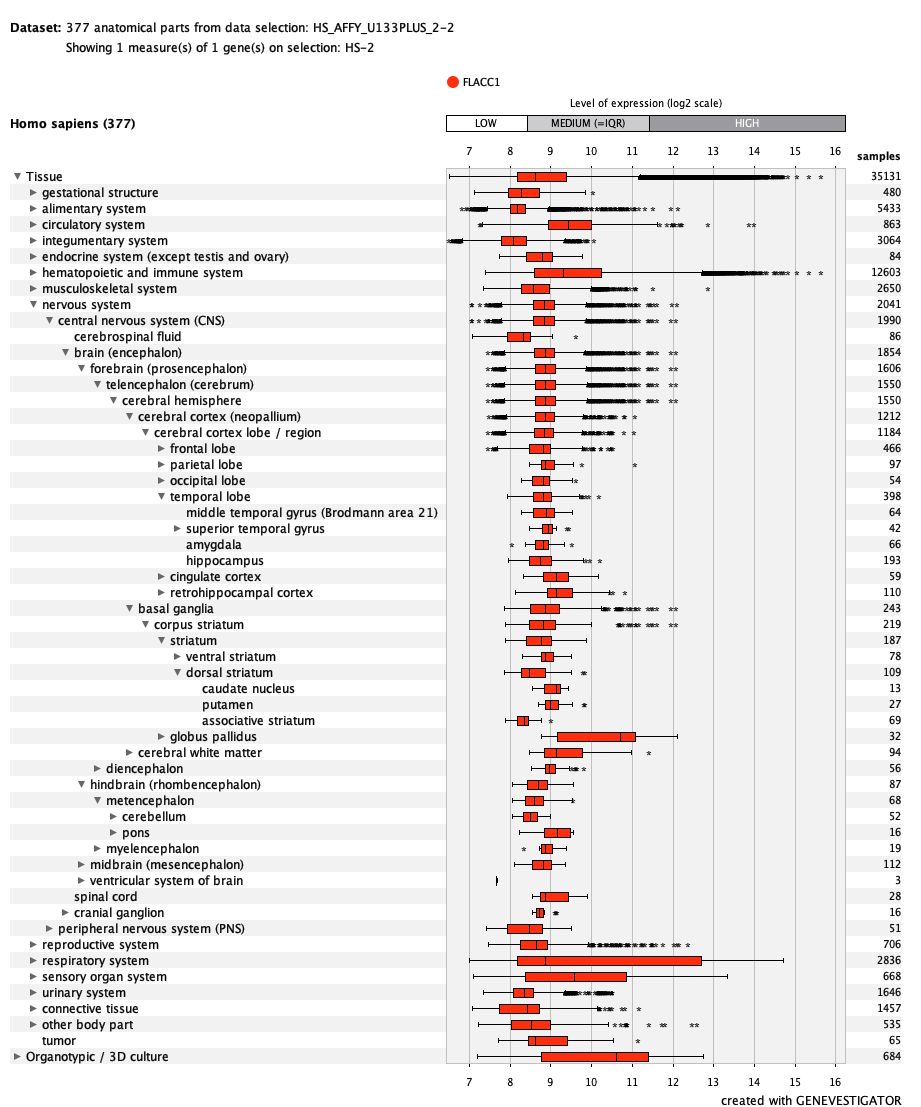

***GCAT***
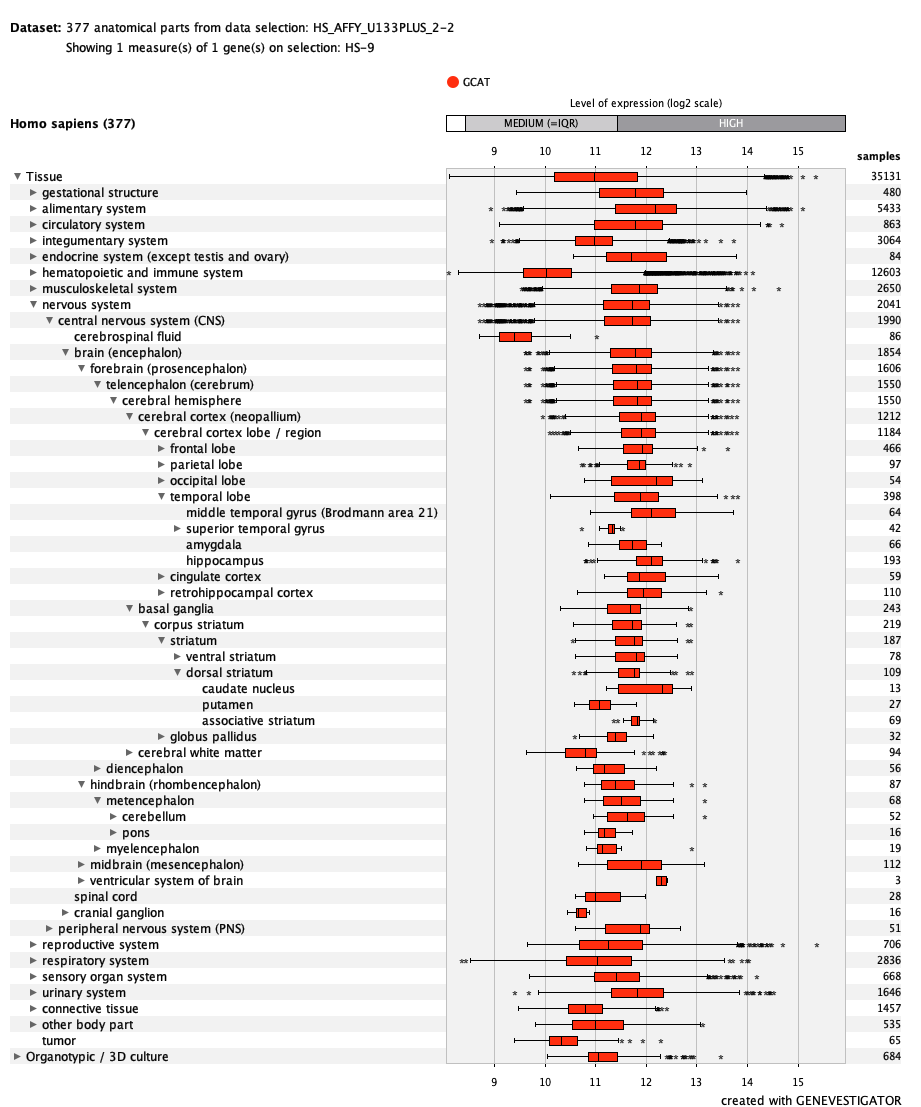

***HIGD1B***
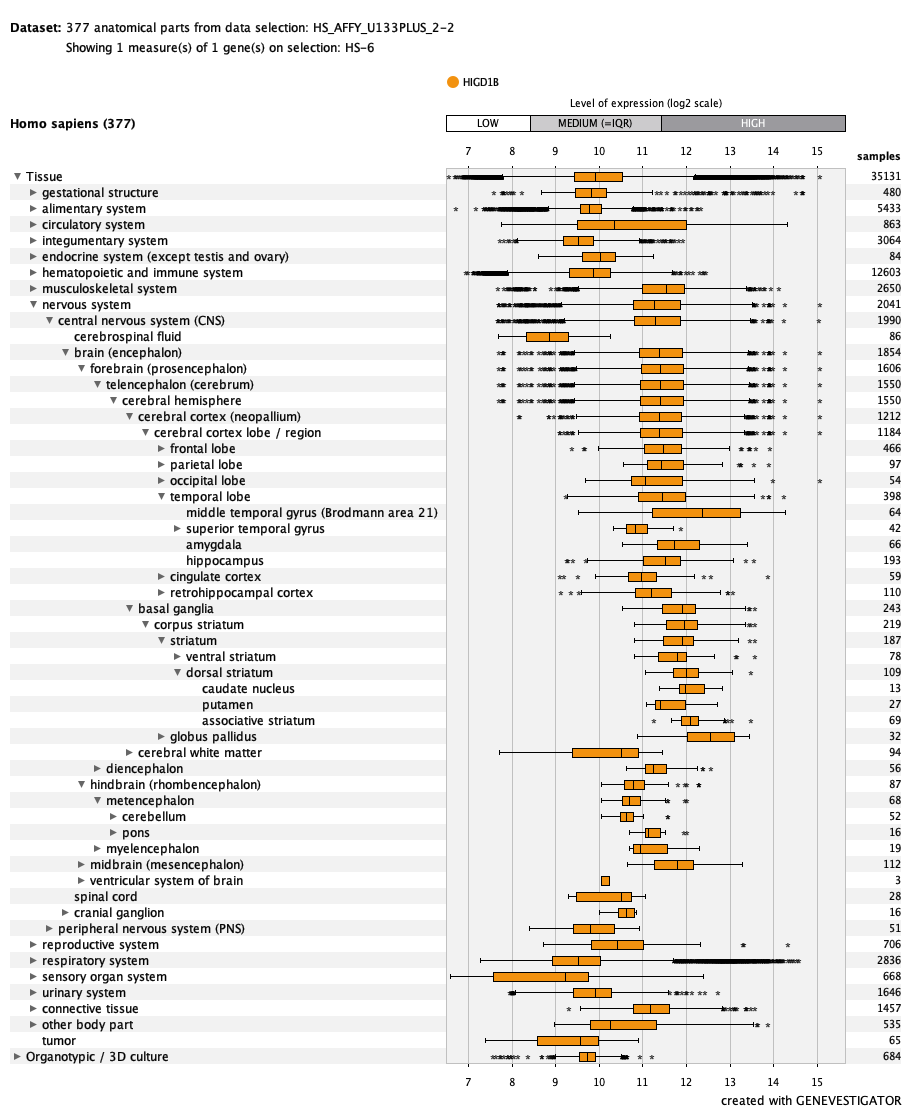

***KANK3***
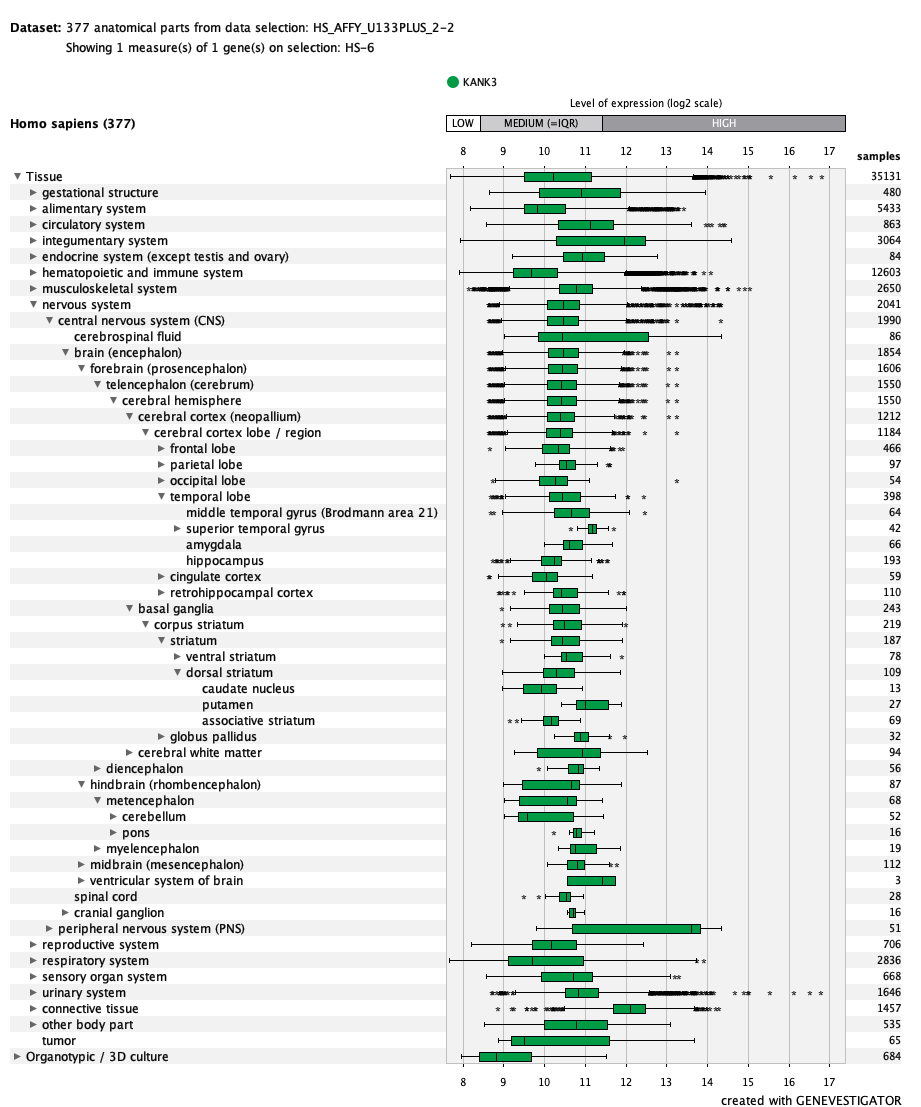

***LLGL2***
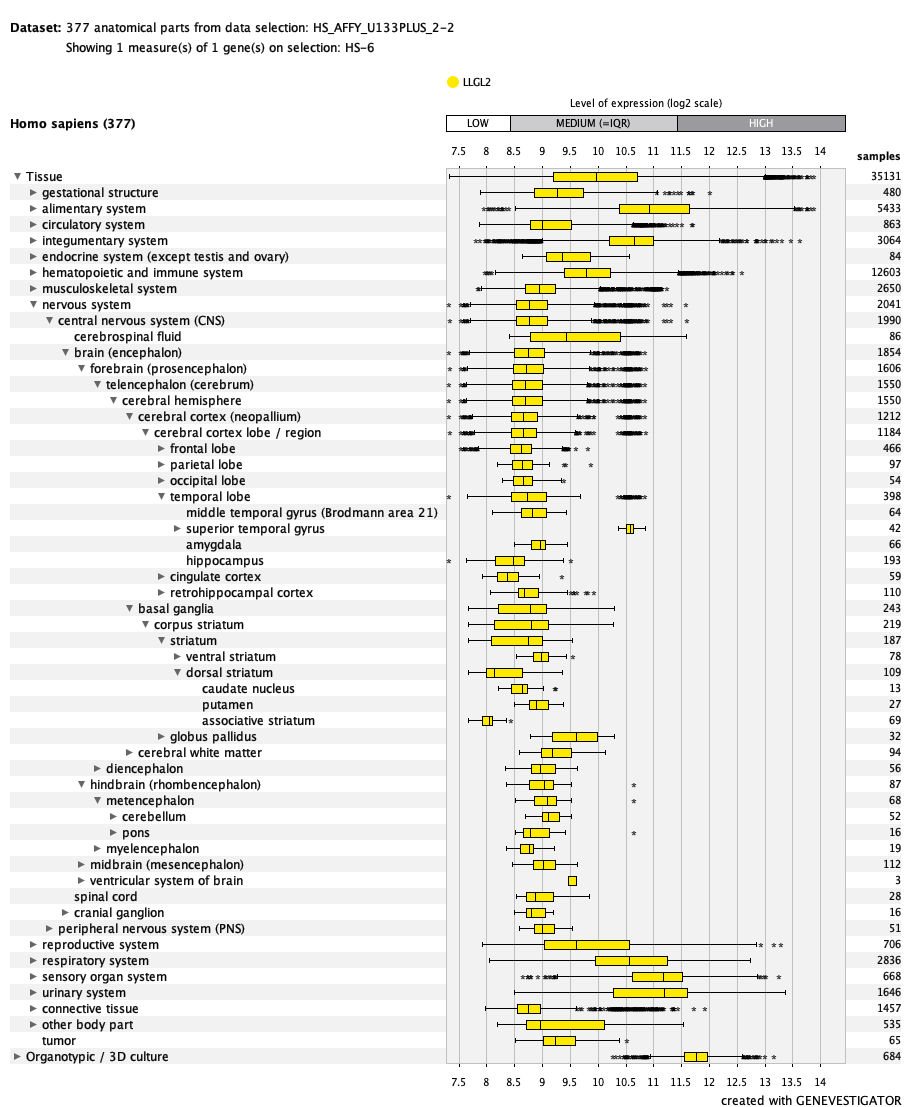

***MMD2***
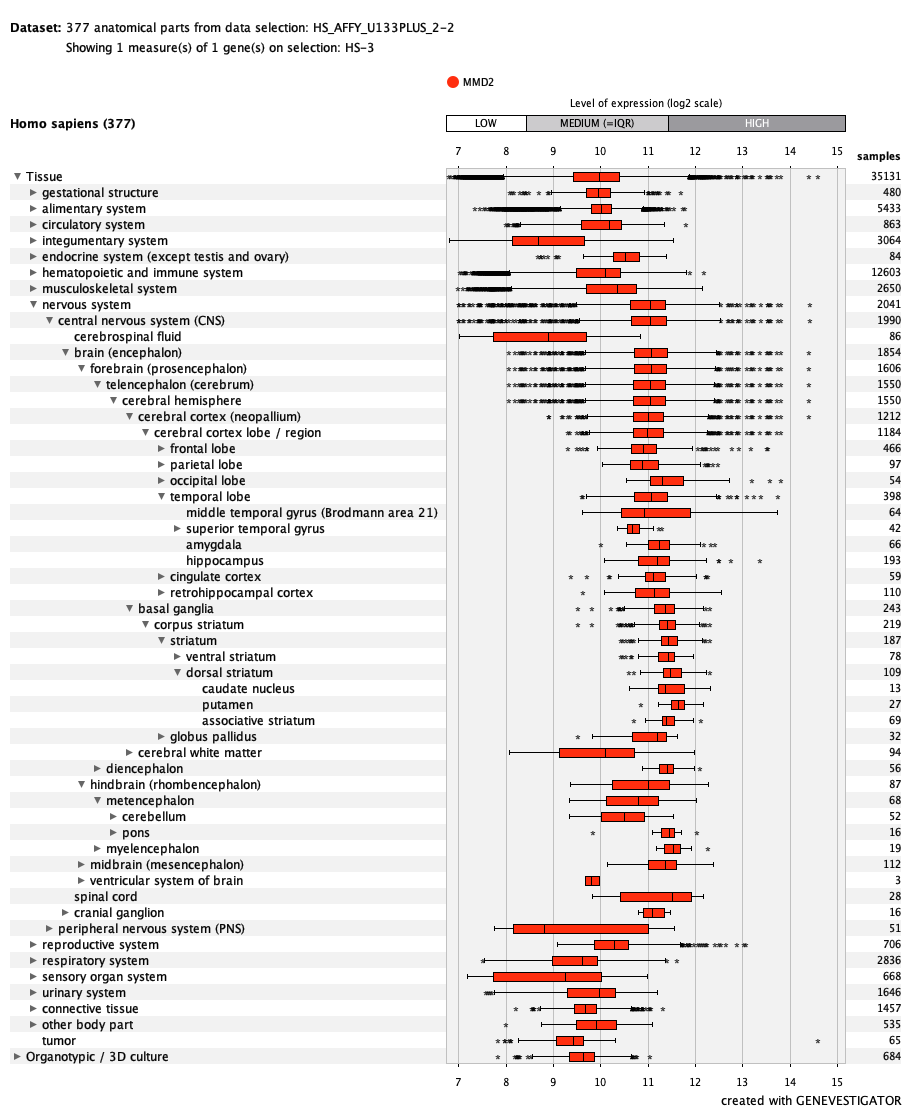

***MMP13***
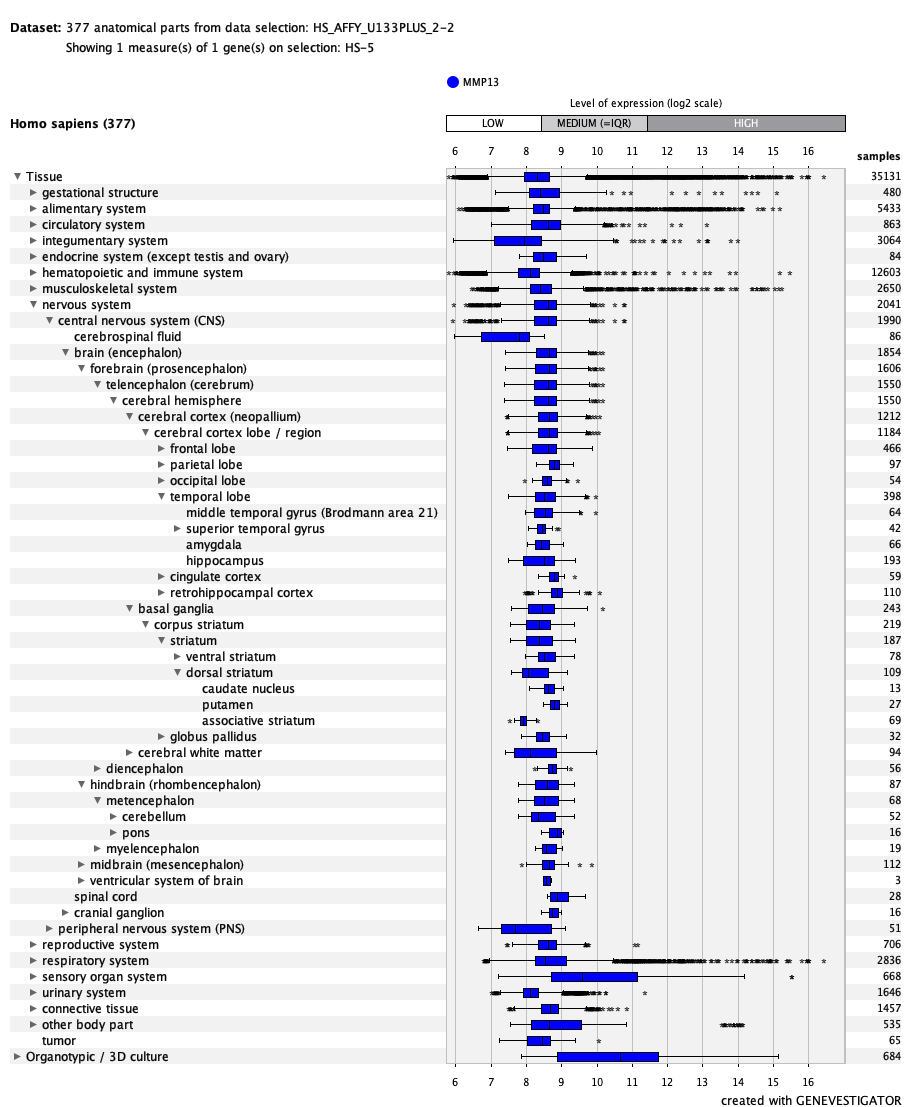

***MMP19***
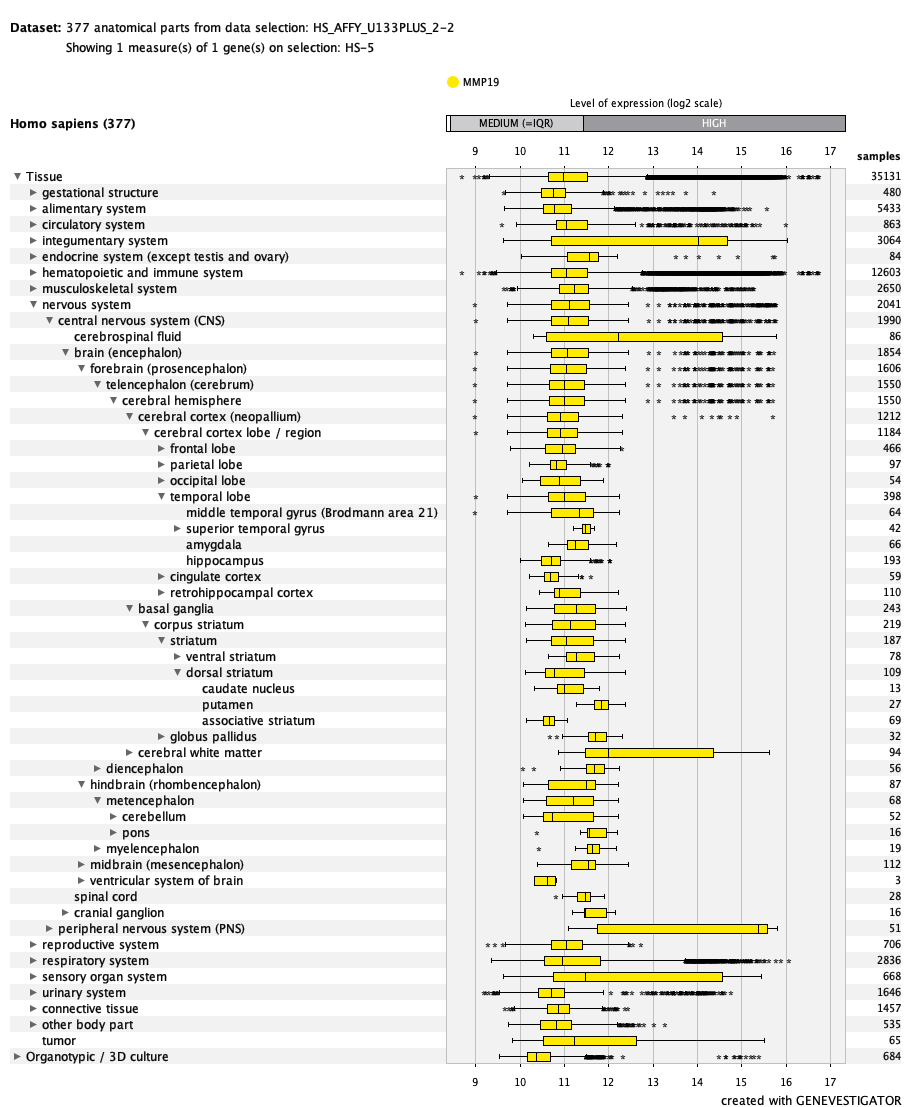

***MUC16***
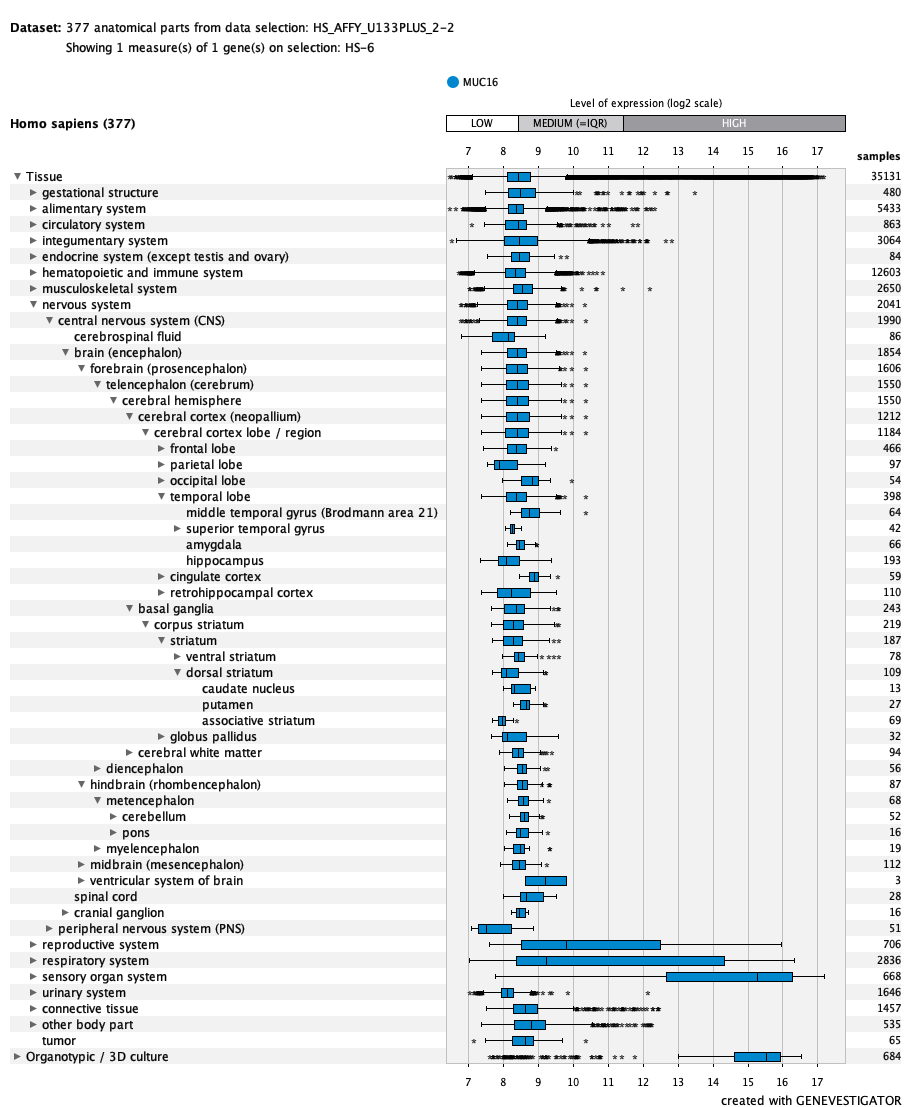

***MUC3A***
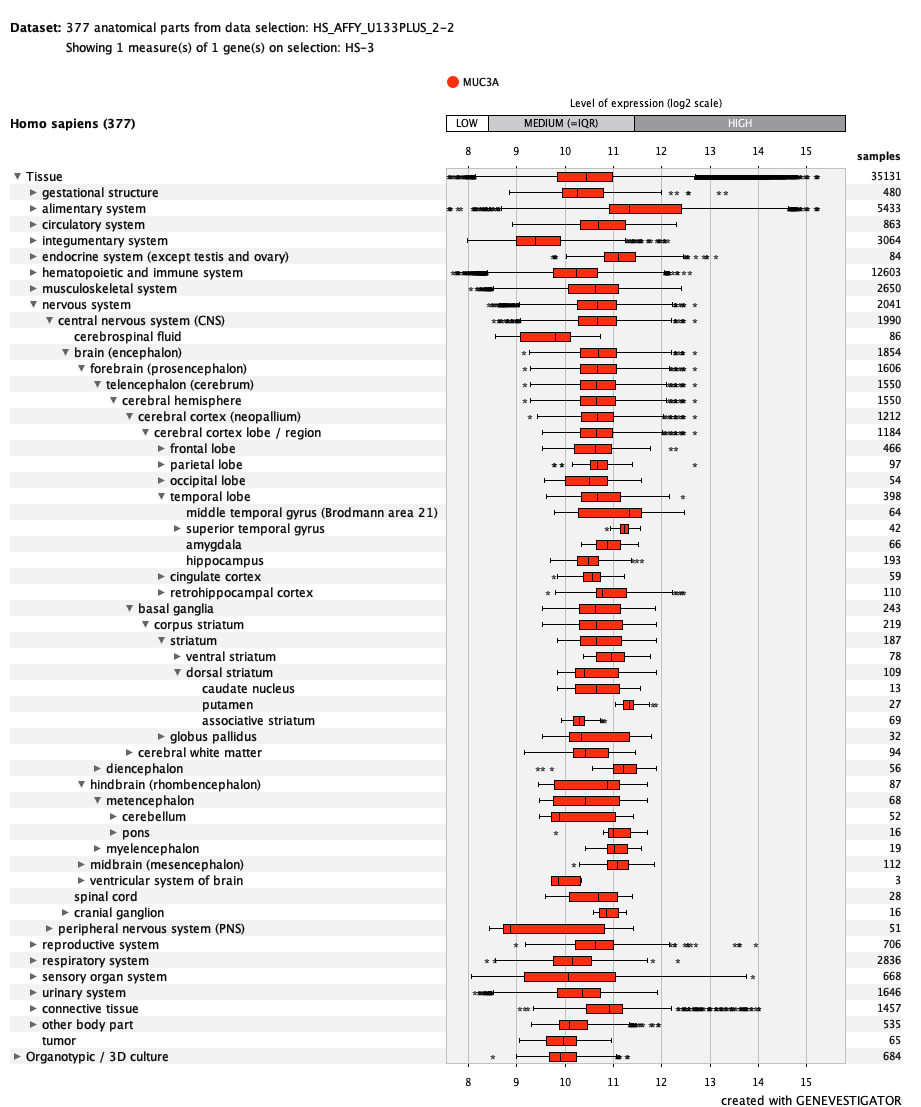

***NOSTRIN***
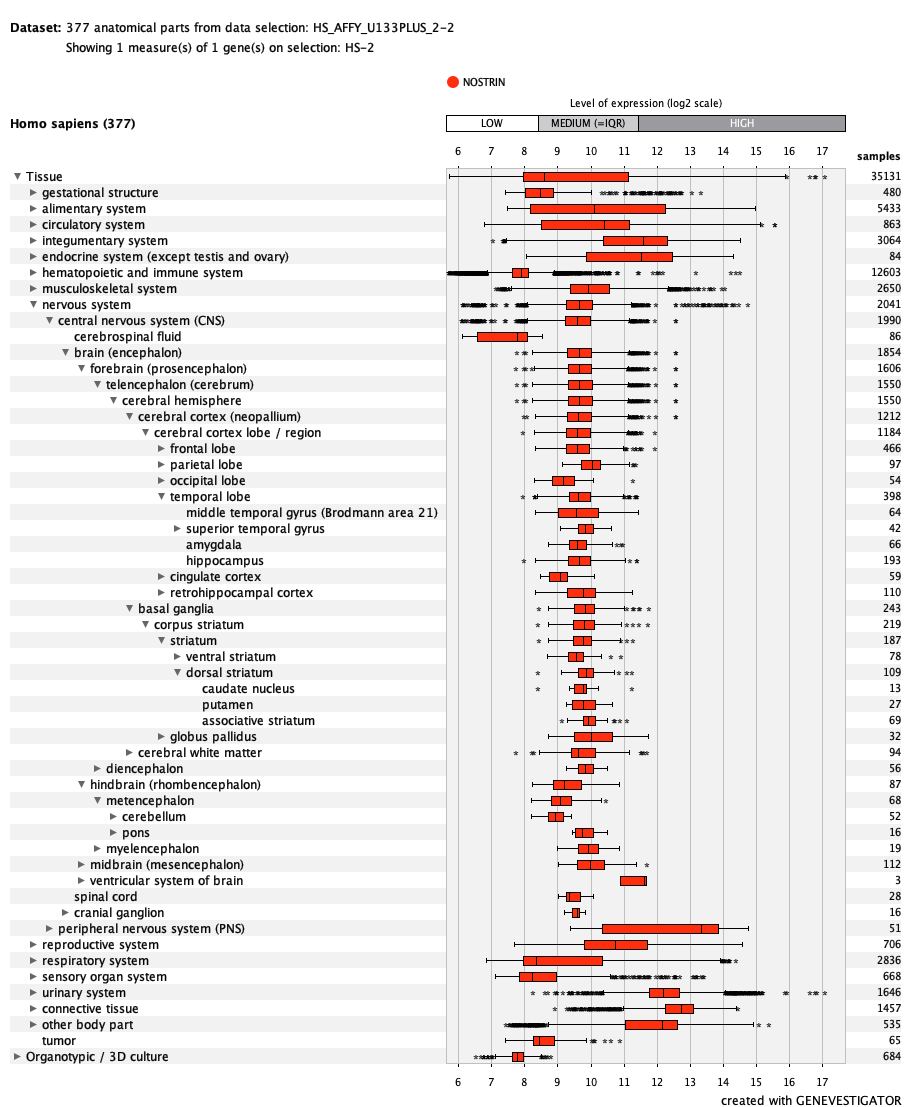

***OMA1***
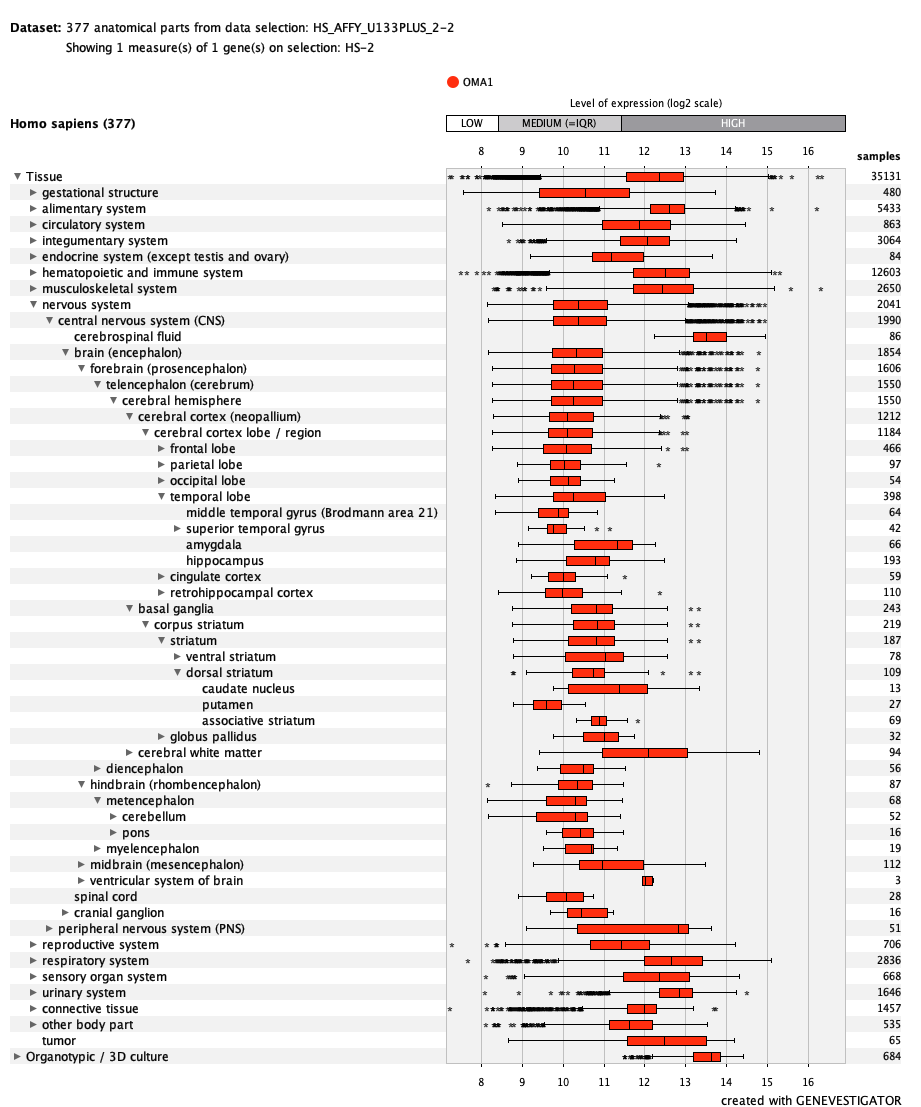

***PPP3CC***
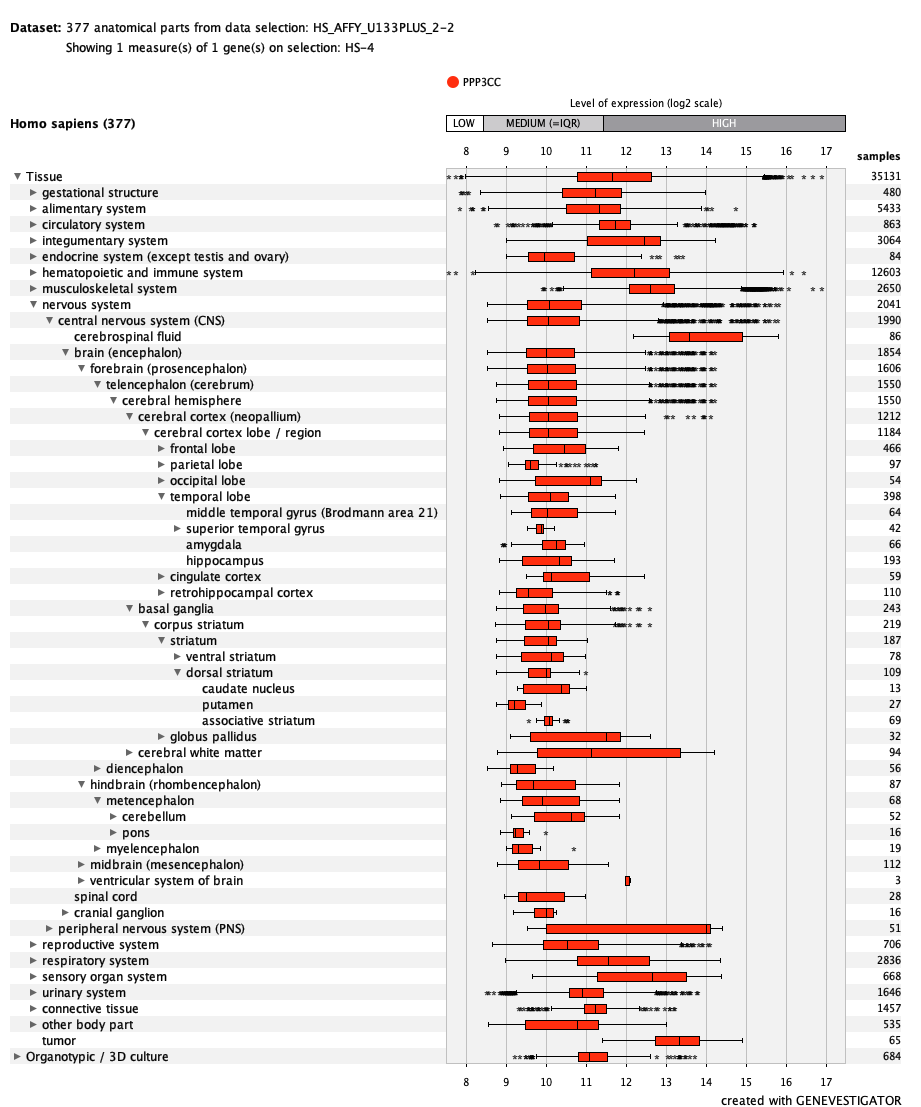

***PRB4***
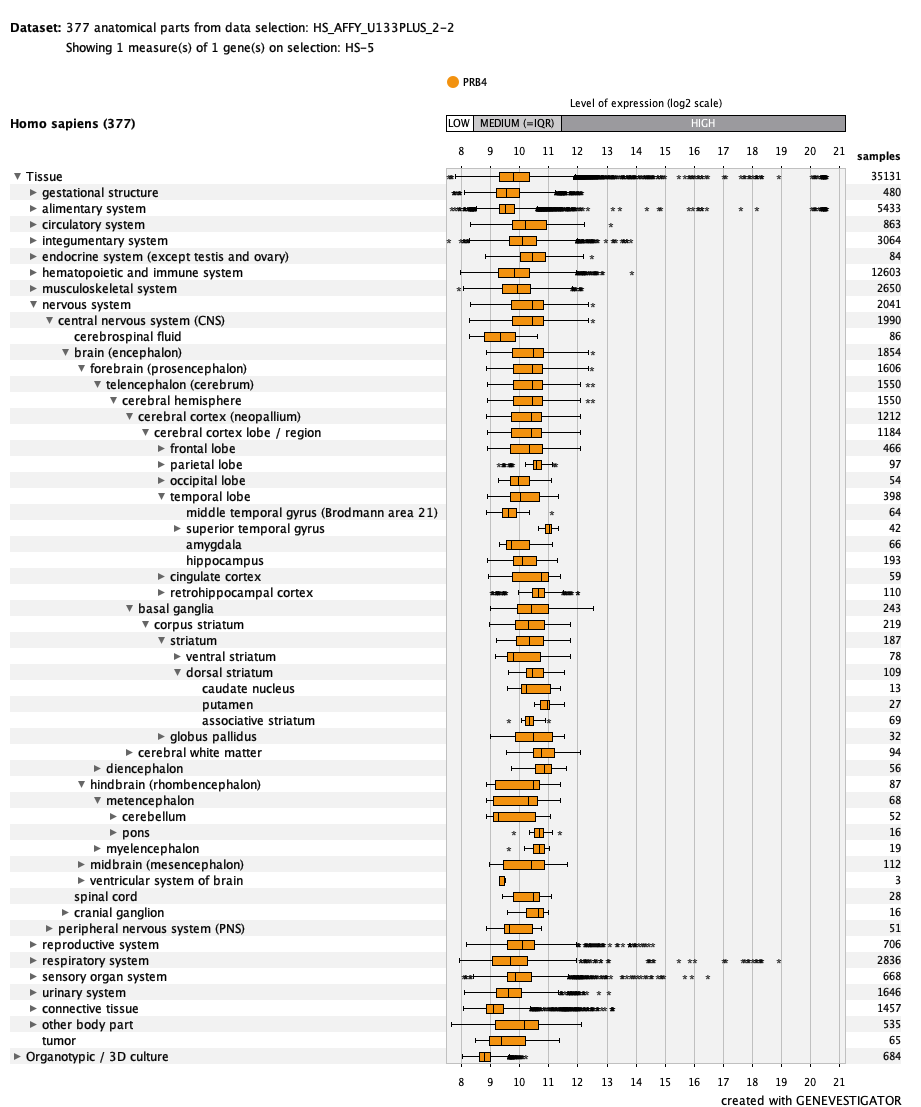

***PRKRA***
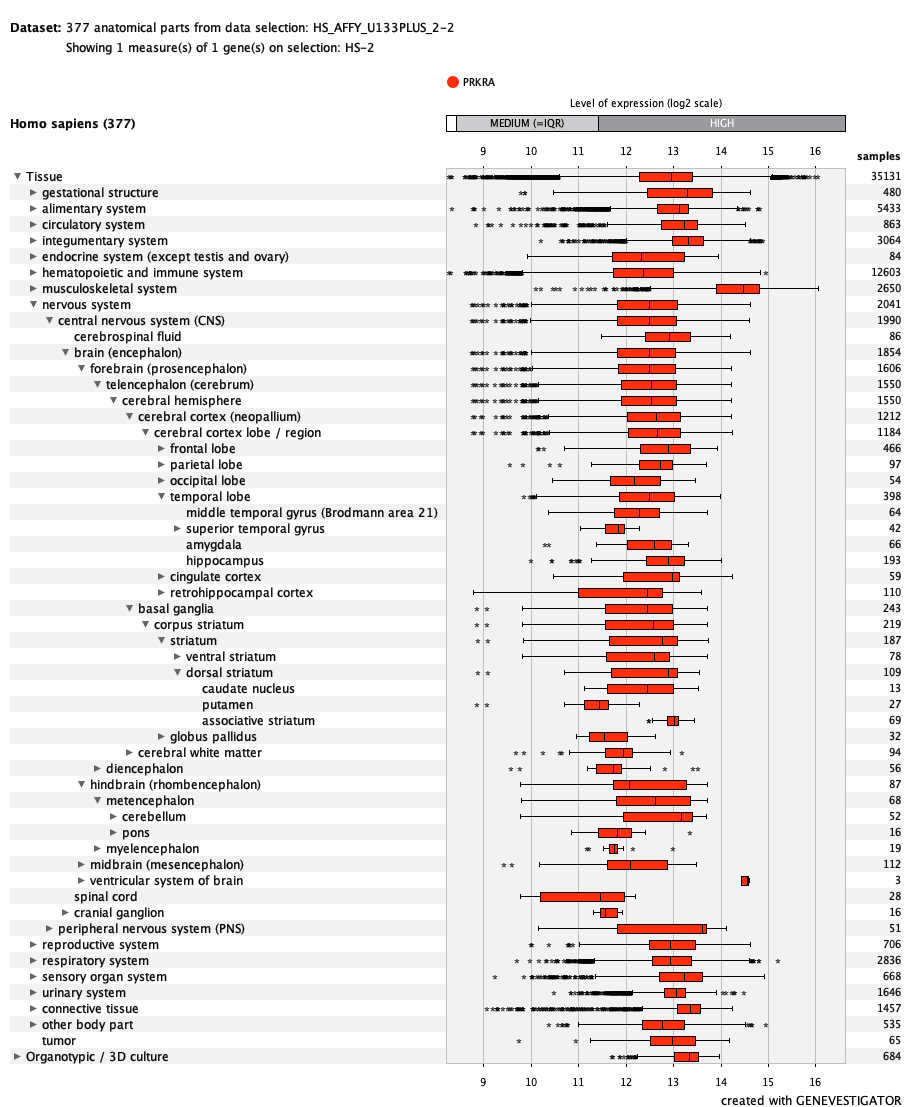

***PRR5***
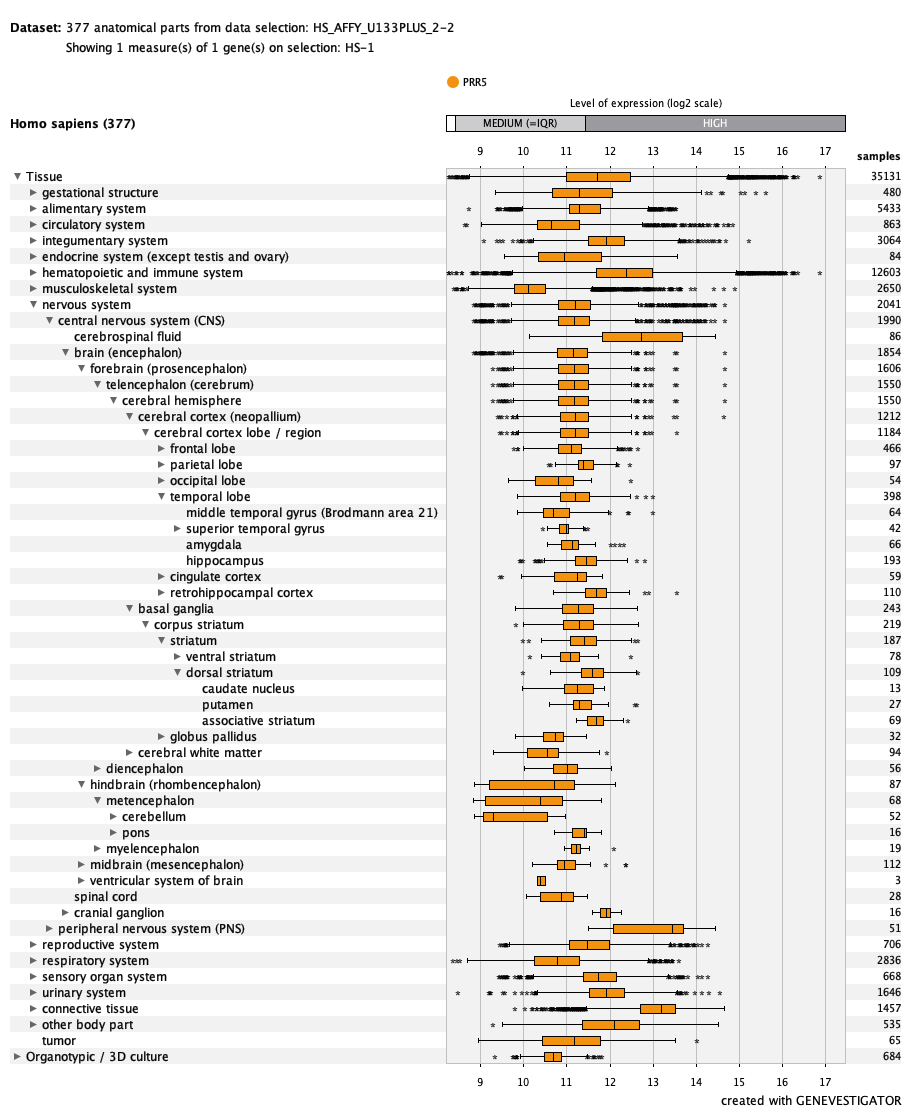

***PRR5-ARHGAP8***
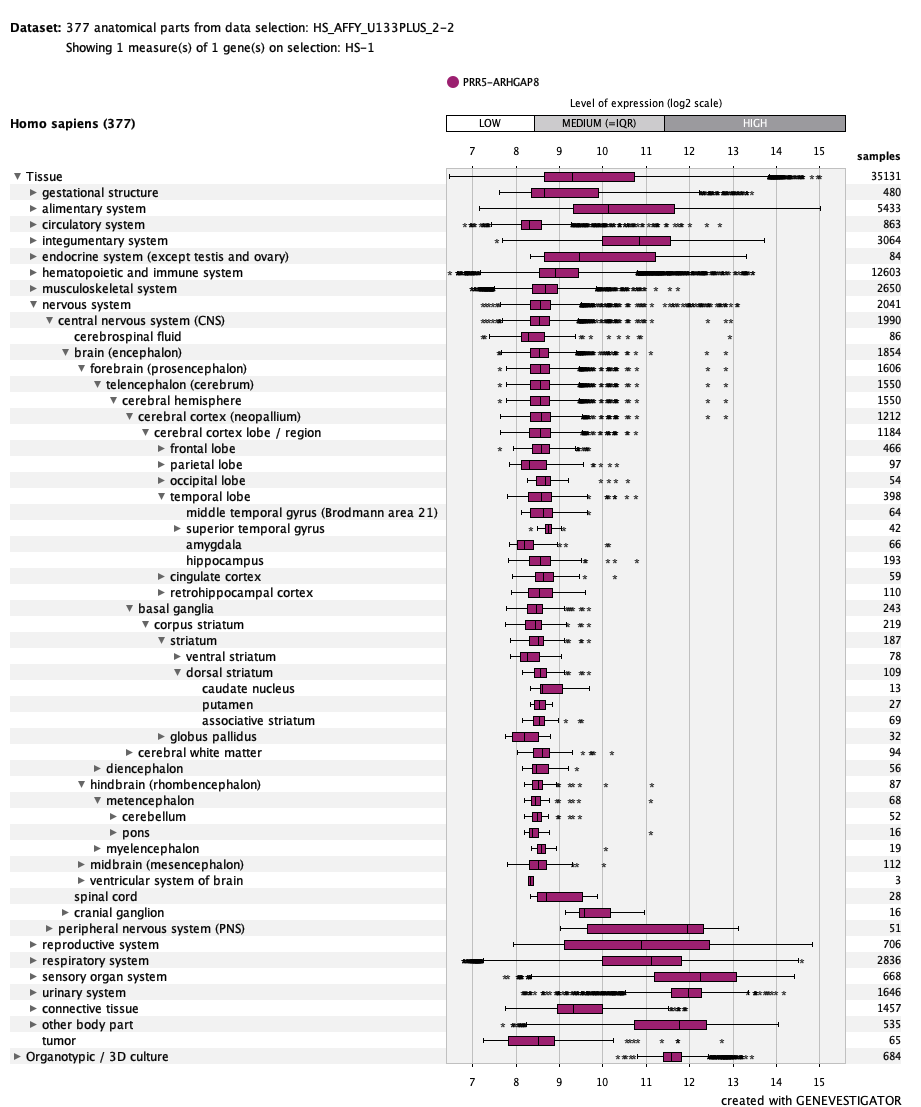

***PRSS3***
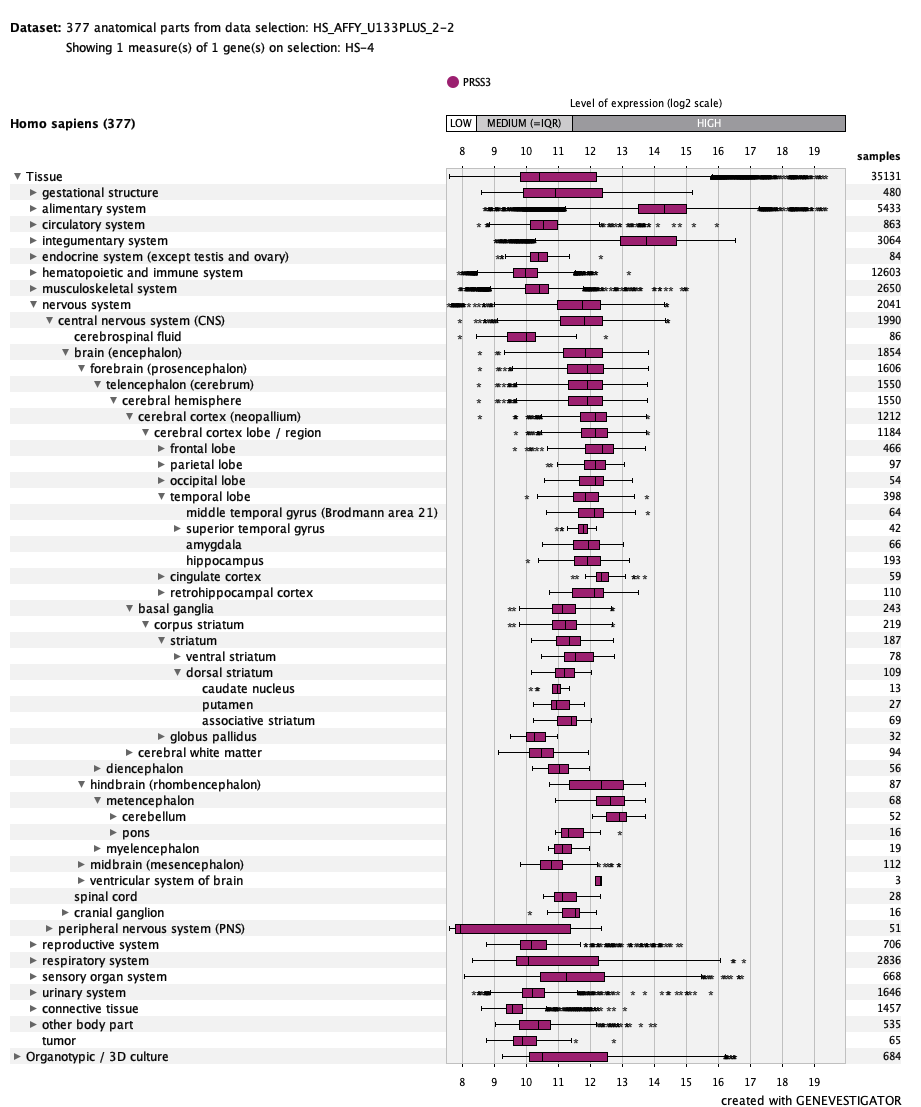

***RASGRF1***
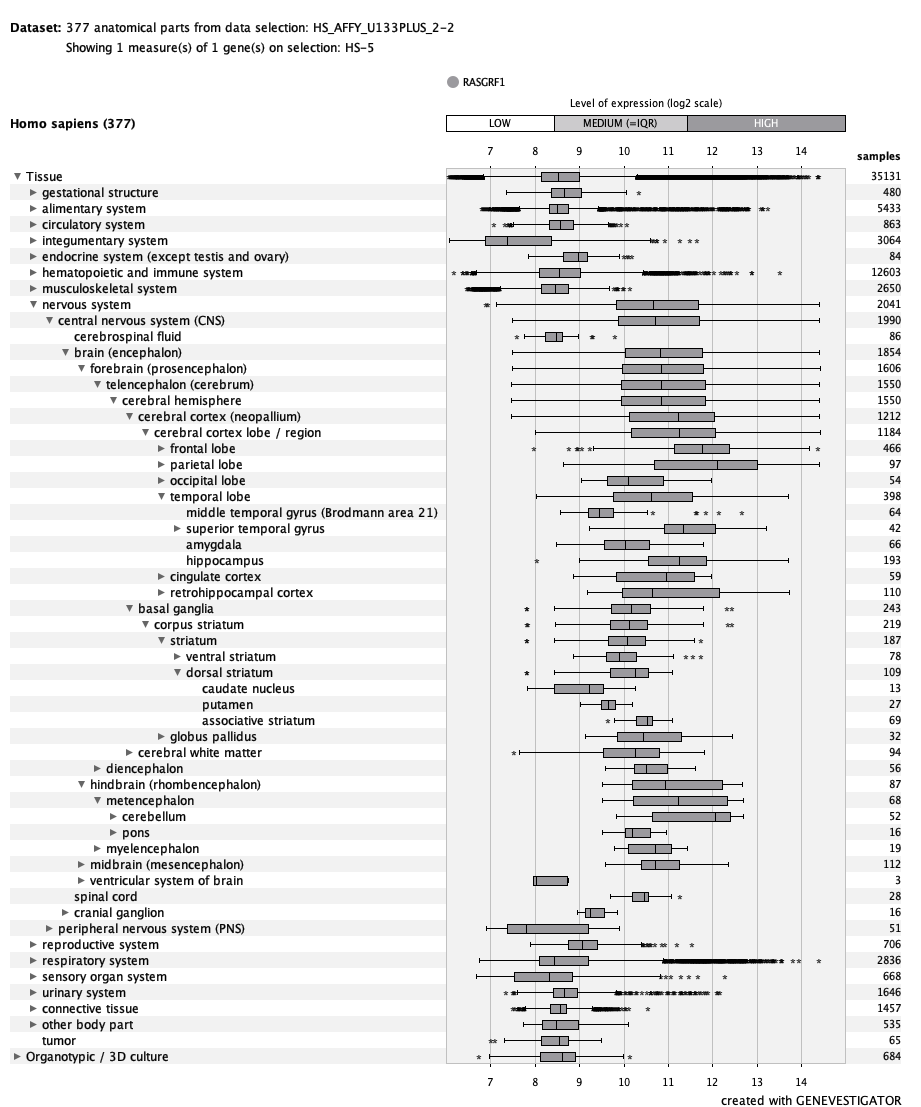

***SFMBT2***
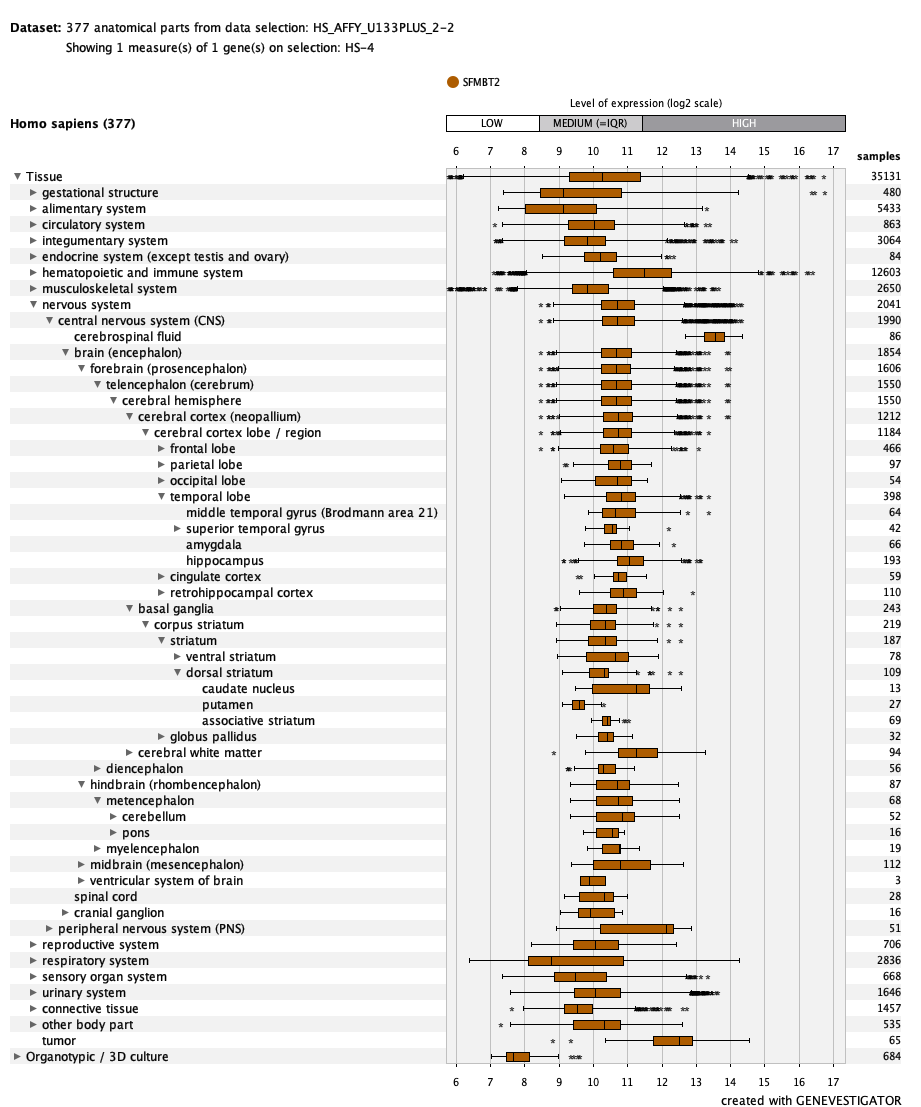

***SPC25***
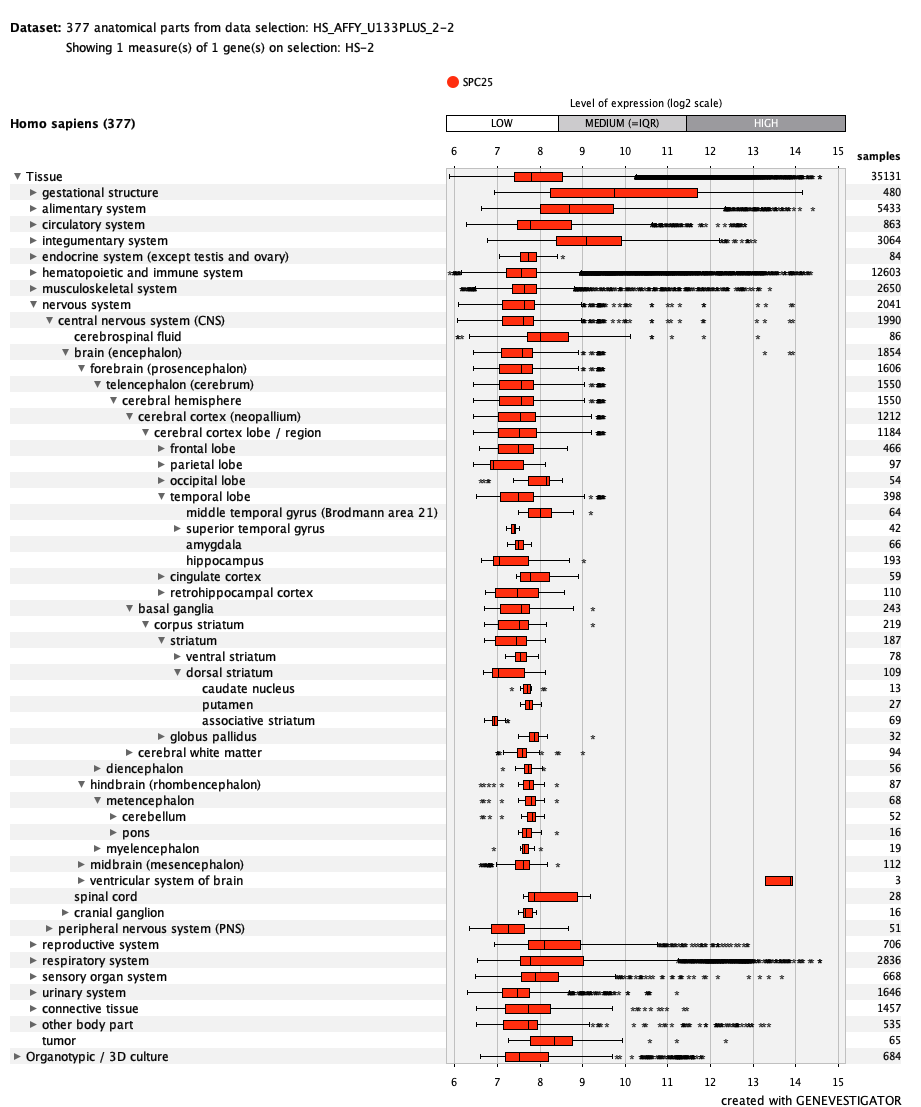

***STK16***
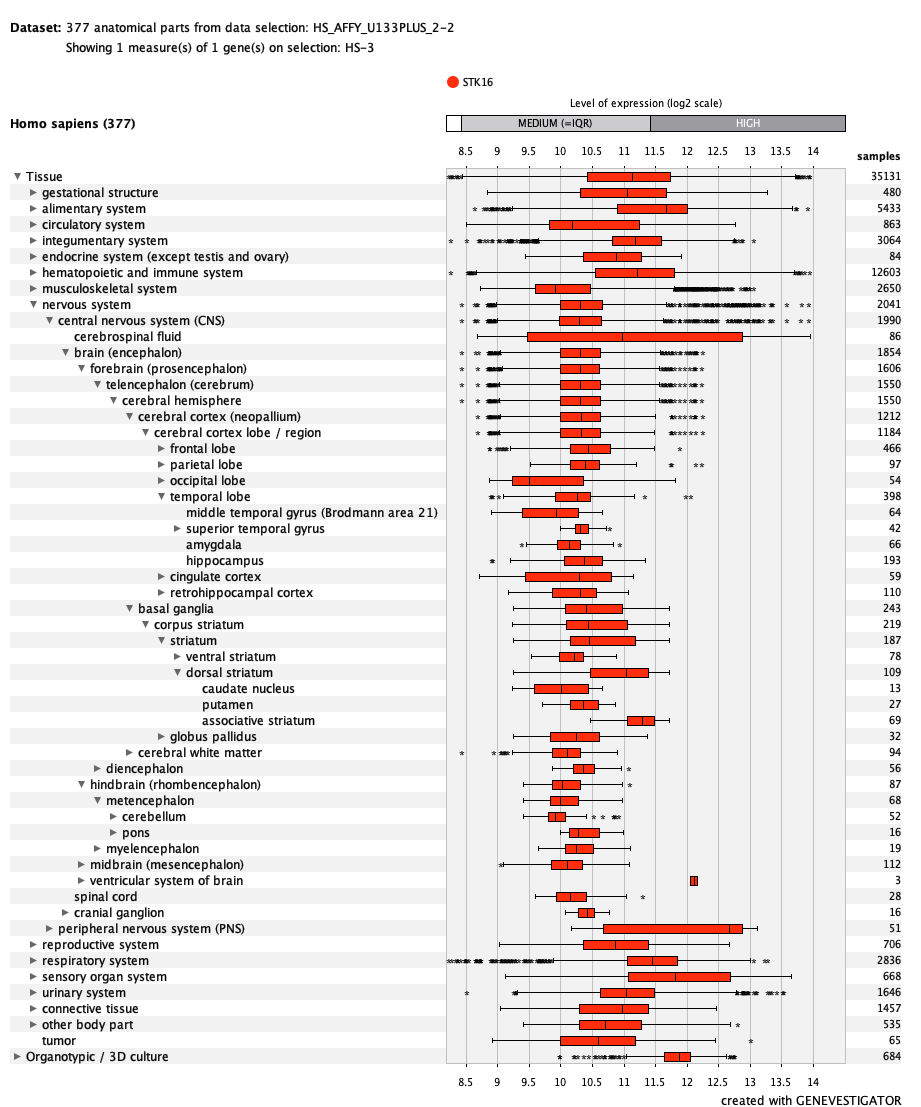

***TTK***
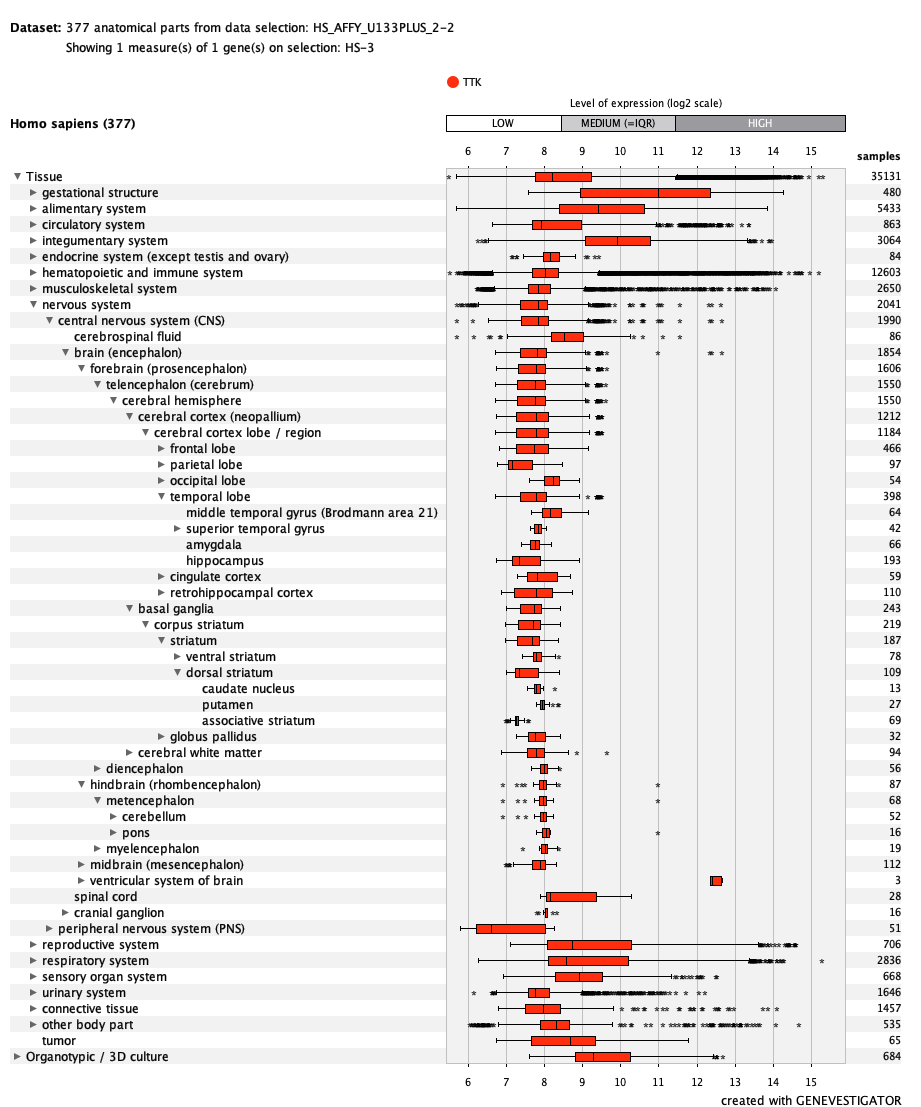

***TUBA4A***
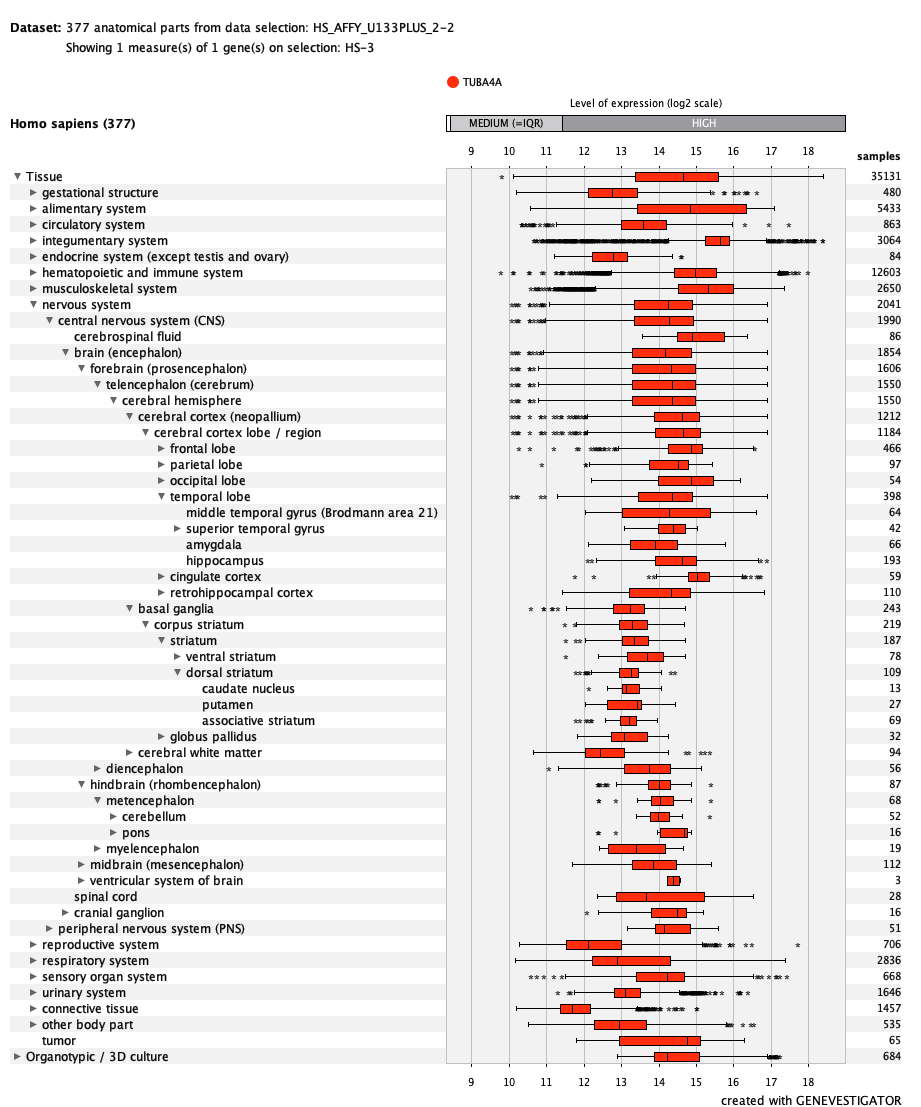

***TUBA4B***
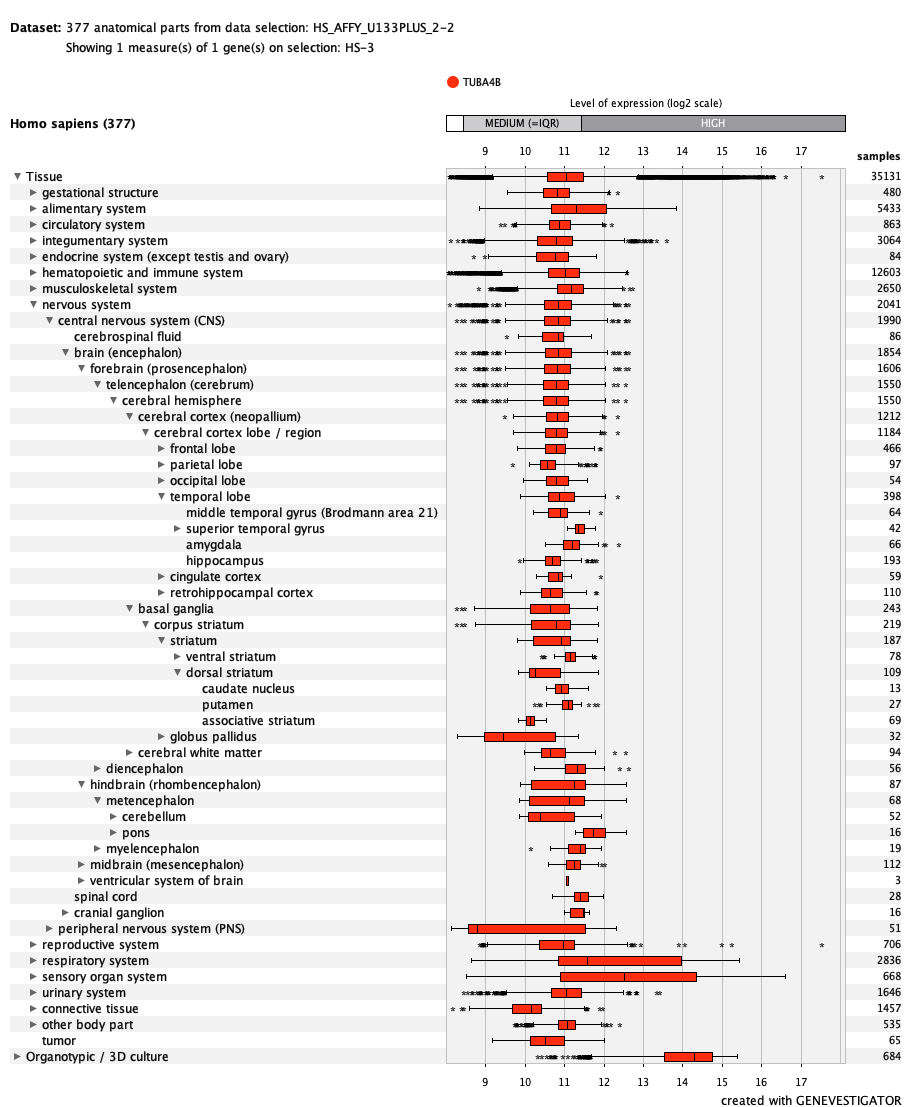

***UBR4***
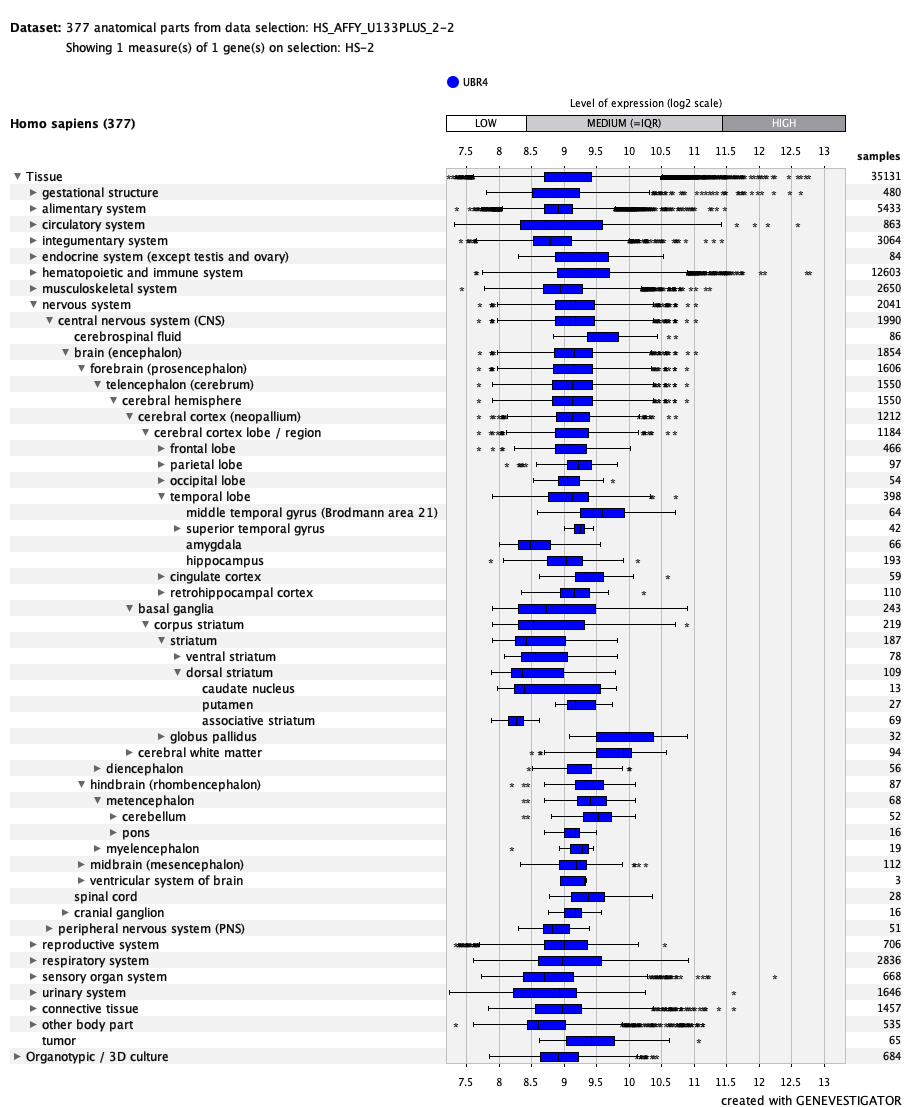

***ZNF296***
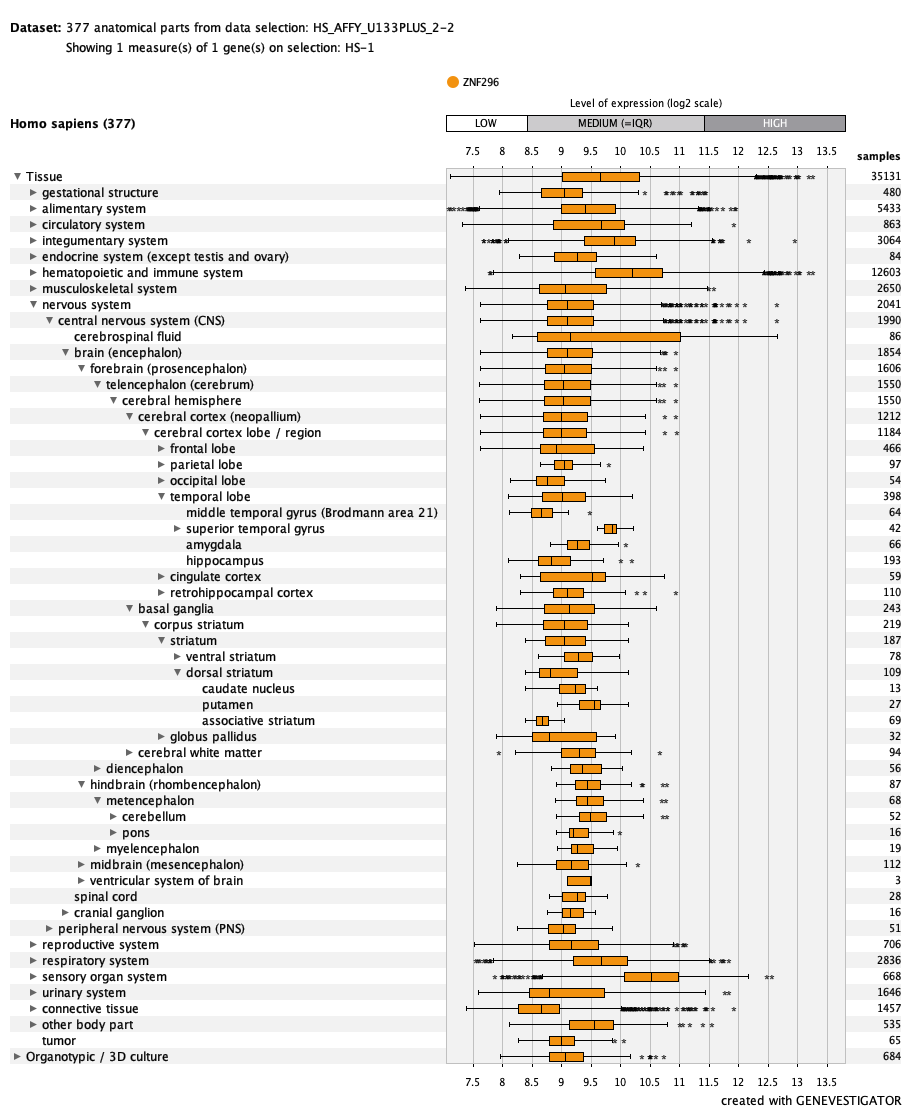
